# Development and implementation of a customised rapid syndromic diagnostic test for severe pneumonia

**DOI:** 10.1101/2020.06.02.20118489

**Authors:** Vilas Navapurkar, Josefin Bartholdson Scott, Mailis Maes, Thomas P Hellyer, Ellen Higginson, Sally Forrest, Joana Pereira-Dias, Surendra Parmar, Emma Heasman-Hunt, Petra Polgarova, Jo Brown, Lissamma Titti, William PW Smith, Jonathan Scott, Anthony Rostron, Matthew Routledge, David Sapsford, M.Estée Török, Ronan McMullan, David A Enoch, Vanessa Wong, The VAPrapid investigators, Martin D Curran, Nicholas M Brown, A John Simpson, Jurgen Herre, Gordon Dougan, Andrew Conway Morris

## Abstract

**Background:** Microbial cultures for the diagnosis of pneumonia take several days to return a result, and are frequently negative, compromising antimicrobial stewardship. The objective of this study was to establish the performance of a syndromic molecular diagnostic approach, using a custom TaqMan array card (TAC) covering 52 respiratory pathogens, and assess its impact on antimicrobial prescribing.

**Methods:** The TAC was validated against a retrospective multi-centre cohort of broncho-alveolar lavage samples. The TAC was assessed prospectively in patients undergoing investigation for suspected pneumonia, with a comparator cohort formed of patients investigated when the TAC laboratory team were unavailable.

Co-primary outcomes were sensitivity compared to conventional microbiology and, for the prospective study, time to result. Metagenomic sequencing was performed to validate findings in prospective samples. Antibiotic free days (AFD) were compared between the study cohort and comparator group.

**Results:** 128 stored samples were tested, with sensitivity of 97% (95% CI 88-100%). Prospectively 95 patients were tested by TAC, with 71 forming the comparator group. TAC returned results 51 hours (IQR 41-69 hours) faster than culture and with sensitivity of 92% (95% CI 83-98%) compared to conventional microbiology. 94% of organisms identified by sequencing were detected by TAC. There was a significant difference in the distribution of AFDs with more AFDs in the TAC group (p=0.02). TAC group were more likely to experience antimicrobial de-escalation (OR 2.9 (95%1.5-5.5).

**Conclusions:** Implementation of a syndromic molecular diagnostic approach to pneumonia led to faster results, with high sensitivity and impact on antibiotic prescribing.

**Trial registration:** The prospective study was registered with clinicaltrials.gov NCT03996330

## Introduction

For many decades the diagnosis of infectious diseases has relied on a combination of clinical assessment and microbiological culture. However, cultures are frequently negative ^1,2^ and can take several days to return a result.^3^ Optimising antimicrobial therapy can be challenging, especially in patients who are at risk of multidrug resistant organisms.^2^ In critically ill patients, this frequently results in the empiric use of broad-spectrum agents, with predictable consequences for antimicrobial resistance and other forms of antimicrobial-related harm.^4^ Conversely, failure to identify the causative organism can lead to inappropriate antimicrobial therapy, which is associated with poor outcomes.^5^

Pneumonia amongst intubated and mechanically ventilated, critically ill patients can be especially difficult to diagnose.^6^ Most critically ill patients are systemically inflamed,^7^ clinical examination is unreliable^8^ and there are multiple causes of radiographic lung infiltrates most of which are non-infectious.^9,10^

The development of host-based biomarkers for infection, such as C-reactive protein,^11^ procalcitonin,^12^ and alveolar cytokine concentrations^13^ have been advanced as useful measures to help rationalise antimicrobial use. However, their utility in the diagnosis^11,12^ and antimicrobial stewardship^14,15^ of pneumonia has been challenged.

There is, therefore, a pressing need for rapid, sensitive, multi-pathogen-focussed diagnostic tests for pneumonia^16^. TaqMan array cards (TAC) enable the conduct of multiple simultaneous single-plex real-time polymerase chain reaction (RT-PCR), with this format allowing rapid and straightforward customisation. Although TACs have shown promising performance relative to conventional microbiology,^17^ our previous experience demonstrated that a TAC with restricted coverage of common respiratory pathogens had a limited impact on clinical decision making in critically ill patients.^18^ We therefore set out to develop and implement a multi-pathogen array that would have broad applicability for severe pneumonia.

## Materials and Methods

### Card development

Organism selection was informed by review the literature concerning organisms found in severe pneumonia ^1,2,6,13,18,19^. Details of organism selection and the card layout are shown in supplemental section (Supplemental Figure S1). The card covers 52 organisms (23 bacteria, 2 mycobacteria, 6 atypical bacteria, 5 fungi and 16 viruses). The study was undertaken prior to the COVID-19 pandemic starting in 2020.

### Card validation

#### Technical validation

The card was initially validated against stored extracts, synthetic control plasmids and all available External Quality Assessment (EQA) panels from Quality Control for Molecular Diagnostics (www.qcmd.org) (supplemental results).

### Retrospective cohort validation

A retrospective cohort validation was conducting using stored bronchoalveolar lavage (BAL) samples obtained during the twenty four centre VAPrapid trial of a biomarker for the diagnosis of ventilator-associated pneumonia.^15^ VAPrapid centres used semi-quantitative microbiological culture as the reference standard.

### Prospective evaluation

#### Setting

Patients were recruited from a 20-bedded teaching hospital Intensive Care Unit (ICU). The unit is a mixed general medical-surgical unit which supports organ and haematology-oncology services.

### Recruitment

Between February 2018 and August 2019, prospectively identified patients were eligible for inclusion if they were receiving invasive mechanical ventilation, and if the treating intensive care specialist was performing diagnostic bronchoscopy for suspected pneumonia. Written consent was obtained from the patient or a proxy decision maker. The TAC laboratory team were routinely unavailable from Friday 5pm to Monday 8am, and also sporadically unavailable due to leave. Patients who were not included in the study because of a lack of TAC laboratory team availability, and those from the month prior and month following the study, formed the comparator group.

### Sampling procedure

Bronchoscopy was undertaken in accordance with existing unit protocols (supplemental methods).

### TAC testing

Nucleic acids were extracted from BAL prior to loading on the TAC. The TAC was run by a dedicated laboratory team who did not undertake the conventional PCR or cultures, with blinding also assured by the results of the TAC being obtained before those from conventional microbiology. Full details of the TAC process are included in the supplemental section.

### Conventional microbiological testing

BAL samples were processed according to the UK Standards for Microbiology Investigations (SMI),^20^ with the results of microbiological semi-quantitative culture and conventional PCR for respiratory viruses, herpesvirade and *Pneumocystis jirovecii* forming the reference standard for the TAC (supplemental methods). As an experimental assay, the results of the TAC were not included in the laboratory information system, blinding the assessors of the reference standard to the TAC results.

### Return of results to clinical team

Following review by a consultant clinical scientist, results were returned to the ICU team. Clinical microbiology advice was available 24 hours/day, and patients underwent weekday daily combined ICU-Microbiology multi-disciplinary review in keeping with existing unit practice (weekend microbiology input was available on request). The study did not mandate any course of action by the treating clinical team. Conventional microbiology results were returned to clinicians via the electronic health record; however in practice these were returned after the TAC results.

### Outcome measures

The co-primary outcome measures were sensitivity, using conventional microbiology as the reference standard and time to result compared to conventional microbial culture. Time to result for microbial culture was taken as time from completion of lavage to first organism identification, or confirmation of negative growth if no organisms were detected.

Secondary outcome measures were sensitivity compared to metagenomic microbial sequencing, time to result compared to conventional PCR, days alive and free of antibiotics (antibiotic-free days, AFDs) in seven and twenty-eight days following lavage and change in antibiotic therapy in the seven days following lavage. Qualitative assessment of whether TAC results impacted on antimicrobial change was assessed by clinical notes review by a member of the study team who was not involved in the decision-making process (VW).

### Statistical analysis

The difference in median time to result for conventional culture and TAC was assessed by Wilcoxon’s matched-pairs test. Where conventional PCR failed, or where the lab did not test for the organism, the corresponding tests from the TAC were removed from calculation of diagnostic performance. Indeterminate cultures (‘mixed upper respiratory tract flora’) were considered negative. A sensitivity analysis, coding failed conventional PCR and organisms not tested ‘negative’ was also undertaken. Comparisons of distribution of antibiotic free days between TAC and comparator groups was by Mann-Whitney U test, differences in proportions of escalation and de-escalation decisions were assessed by Chi^2^ test. Analyses were conducted using Prism v9.1 (Graphpad Inc, La Jolla, CA).

### Study size

A planned prospective study size of 100 patients evaluated by TAC was selected to balance cost against including sufficient numbers to be able to make a judgement on the card’s clinical utility. As the co-primary endpoint was time to result in a real-world setting that had not been previously evaluated, we did not undertake a formal power calculation.

### Ethical and regulatory approvals and funding

The prospective study was approved by the Leeds East Research Ethics Committee (17/YH/0286) Cambridge University Hospitals NHS Foundation Trust was the sponsor, and registered with clinicaltrials.gov (NCT03996330). The assessment of routinely collected data from the comparator group received a consent waiver and was conducted under a protocol approved by the institutional review board (A095506). VAPrapid^15^ was approved by the England and Northern Ireland (13/LO/065) and Scotland (13/SS/0074) National Research Ethics Service committees and sponsored by Newcastle upon Tyne Hospitals NHS Foundation Trust.

## Results

### Technical validation

Following initial validation against stored DNA extracts and synthetic plasmids, all microorganisms from the Quality Control for Molecular Diagnostics 2018 Sepsis EQA Pilot Study were successfully detected (supplemental Table S1).

### Retrospective cohort validation

The card was tested against the stored samples available from the VAPrapid study^15^. 128 samples with semi-quantitative culture results were available for analysis. 57 organisms were grown at or above 10^4^ colony forming units(CFU)/ml ^20,21^, with 55 detected by TAC (Table 1). The TAC detected a further 295 organisms, including 64 viruses and one atypical organism which the recruiting centres did not test for. Excluding tests for organisms not detectable by culture, 3425 tests on TAC were negative. Sensitivity was 97% (95% CI 88-100%) and specificity 94% (95% CI 93-95%) (Supplemental Table S2). Organisms detected by both TAC and culture had a median cycles to threshold (Ct) value on the TAC of 29 (IQR 26-32 range 20-35) whilst those detected by TAC alone had a median Ct value of 33 (IQR 30-35 range 20-40) (supplemental Figure S2).

**Table 1:** culture of microorganisms from 128 stored samples from the VAPrapid clinical trial^15^ and results from the TAC. CFU-colony forming units/ml, Ct -cycles to crossing threshold. *not on card ^#^ culture reported as *Proteus mirabilus*, on TAC reported as genus-level *Proteus spp.*

| Organism detected | Frequency of growth ($\geq 10^4$ CFU/ml) on conventional culture | Frequency by TAC (numbers detected at $Ct \leq 32$ shown in brackets) |
| --- | --- | --- |
| **Gram negative** |  |  |
| *Acinetobacter baumannii* complex | 2 | 4 (3) |
| *Enterobacter aerogenes* | 1 | 0* |
| *Enterobacter cloacae* | 2 | 7 (5) |
| *Enterococcus faecalis* | 0 | 2 (1) |
| *Enterococcus faecium* | 2 | 15 (9) |
| *Escherichia coli* | 6 | 44 (16) |
| *Enterobacteriaceae* (not further specified) | 0 | 7 (3) |
| *Haemophilus influenzae* | 3 | 23 (19) |
| *Haemophilus haemolyticus* | 1 | 0* |
| *Klebsiella pneumoniae* | 2 | 13 (6) |
| *Legionella* spp. (non-pneumophilia) | 0 | 1 (1) |
| *Moraxella catharralis* | 1 | 5 (4) |
| *Morganella morganii* | 0 | 1 (1) |
| *Mycoplasma pneumoniae* | 0 | 1 (1) |
| *Neisseria meningitidis* | 0 | 1 (1) |
| *Proteus* spp.<sup>#</sup> | 2 | 7 (5) |
| *Pseudomonas aeruginosa* | 5 | 10 (9) |
| *Serratia marcescens* | 1 | 5 (3) |
| **Gram positive** |  |  |
| *Staphylococcus aureus* | 21 | 32 (28) |
| *Staphylococcus epidermidis* | 0 | 12 (3) |
| Other coagulase negative *Staphylococcus* | 0 | 9 (1) |
| *Stenotrophomonas maltophilia* | 2 | 11 (5) |
| *Streptococcus pneumoniae* | 0 | 15 (10) |
| *Streptococcus pyogenes* | 0 | 1 (1) |
| *Streptococcus* spp. (not further specified) | 0 | 37 (25) |
| **Fungi** |  |  |
| *Aspergillus fumigatus* | 0 | 1 (1) |
| *Candida albicans* | 4 | 17 (7) |
| *Candida* spp. | 2 | 5 (3) |
| **Viruses** (not tested for by conventional microbiology) |  |  |
| Coronavirus OC43 |  | 1 (1) |
| Cytomegalovirus |  | 6 (1) |
| Epstein-Barr Virus |  | 15 (3) |
| Herpes simplex virus |  | 34 (26) |
| Influenza A |  | 3 (3) |
| Parainfluenza virus |  | 1 (1) |
| Rhinovirus |  | 4 (4) |

### Prospective evaluation

Between January 2018 and September 2019 166 ventilated patients were investigated for pneumonia by bronchoscopy, 95 were tested by TAC. Five patients were tested twice by TAC, having suffered a subsequent respiratory deterioration, in total 100 TACs were run. 71 patients formed the comparator group (Figure 1). Although inclusion criteria were pragmatic and only required senior clinician suspicion of pneumonia, 92% of cases met full ECDC criteria for clinical pneumonia (Supplemental Figure S3). Of the eight cases not meeting full ECDC criteria, one lacked a formal radiological report of infiltrates, one had no clinical signs of pneumonia, five had no signs of systemic inflammation and one patient lacked both radiological and systemic inflammation. Table 2 shows participant characteristics of the study population and comparator group.

**Table 2:** Baseline characteristics of study population. APACHE II, acute physiology and chronic health evaluation II, FiO_2_, fraction of inspired oxygen.

| Parameter | TAC patients<br>(95 patients) | Comparator group<br>(71 patients) |
| --- | --- | --- |
| Median age (range) | 60 (21-86) | 62 (18-83) |
| n (%) female | 41 (43 %) | 27 (38 %) |
| Median (IQR) functional<br>comorbidity index | 1 (2) | 1 (2) |
| n (%) with community-<br>acquired pneumonia | 34 (36%) | 20 (28%) |
| n (%) hospital-acquired<br>pneumonia | 61 (64%) | 51 (72%) |
| of HAP n (%) ventilator-associated | 24 (39%) | 27 (52%) |
| Median (IQR) APACHE II<br>score on admission | 16 (10) | 16 (9) |
| % receiving antibiotics at time<br>of lavage | 82% | 96% |
| Median (IQR) FiO<sub>2</sub> prior to<br>bronchoscopy | 0.5 (0.25) | 0.5 (0.30) |
| Median (IQR) white cell count<br>(x10<sup>9</sup>/L) | 10.5 (12.4) | 10.7 (9.5) |
| Median (IQR) neutrophil count<br>(x10<sup>9</sup>/L) | 8.6 (11.5) | 8.8 (8.63) |
| Median (IQR) C-reactive<br>protein concentration<br>(mg/L) | 198 (153) | 146 (154) |
| 28-day mortality n (%) | 30 (32%) | 21 (30%) |

**Figure 1:**
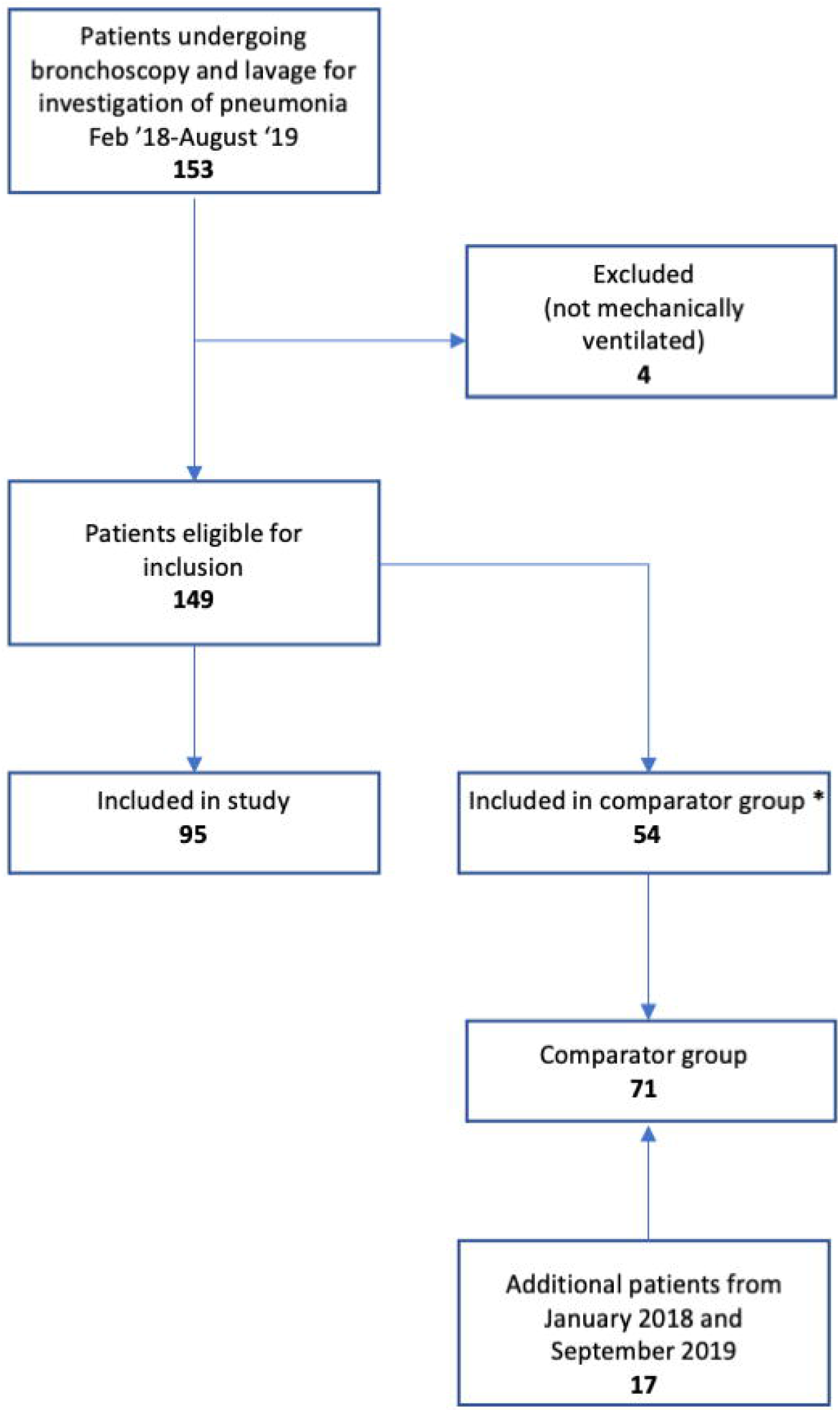
Study flow diagram. *included in comparator group as TAC laboratory team not available to process samples

**Figure 2:**
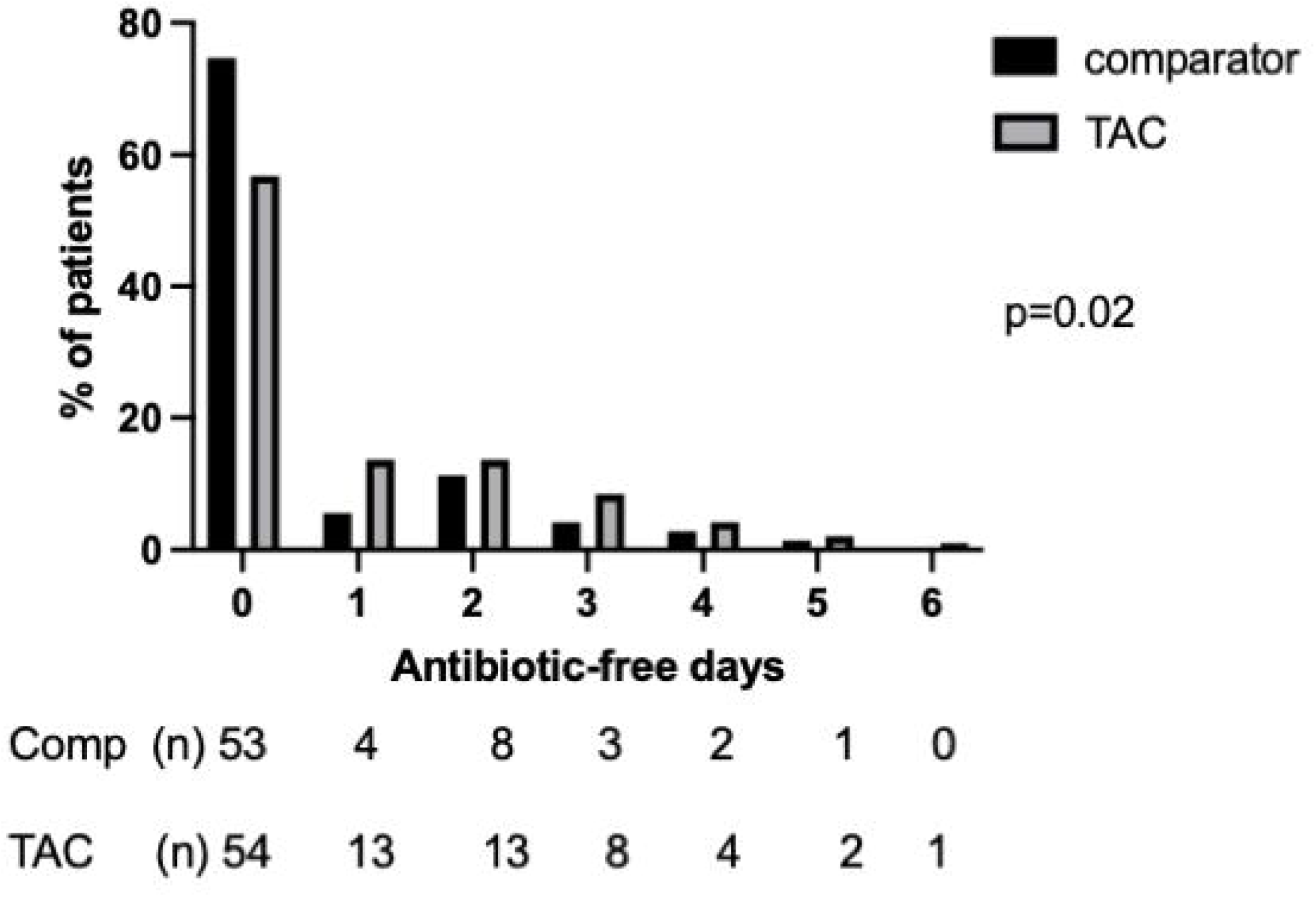
Distribution of days alive and free of antibiotics in the seven days following bronchoscopy and lavage in the TAC and comparator cohorts. Following first lavage only for patients who had more than one BAL during ICU admission. Numbers in each category and percentage shown below graph, p value by Mann-Whitney U test.

### Time to result

The median difference in time to result between TAC and conventional culture was 51 hours (IQR 41-69 hours p<0.0001 by Wilcoxon matched pairs), the TAC also returned results more rapidly than conventional PCR in almost all cases (Supplemental Figure S4). The minimum TAC time to return was 4 hours, with median time to result 22 hours (IQR 7-24 hours), most of the delays arose from samples taken outside routine working hours, whilst additional delays with conventional PCR results largely reflect laboratory workflow and batching of samples.

### Comparison of organisms detected by TAC compared to conventional microbiology

178 organisms were identified from 100 samples on the TAC (Table 3, Supplemental Table S3). Conventional microbiology detected 66 organisms, with 61 detected by TAC. 27 patients had failure of internal control for one or more conventional PCR assays, covering 93 organisms. There were no TAC internal control failures and none of the organisms covered by the failed assays were detected on TAC or sequencing (Table 3). Sensitivity and specificity were 92% (95% CI 83-98%) and 97% (95% CI 97-98%) respectively (Supplemental Table S4). Including failed and absent reference standards as ‘negative’ had minimal effect on diagnostic performance (Supplemental Table S5)

**Table 3:**
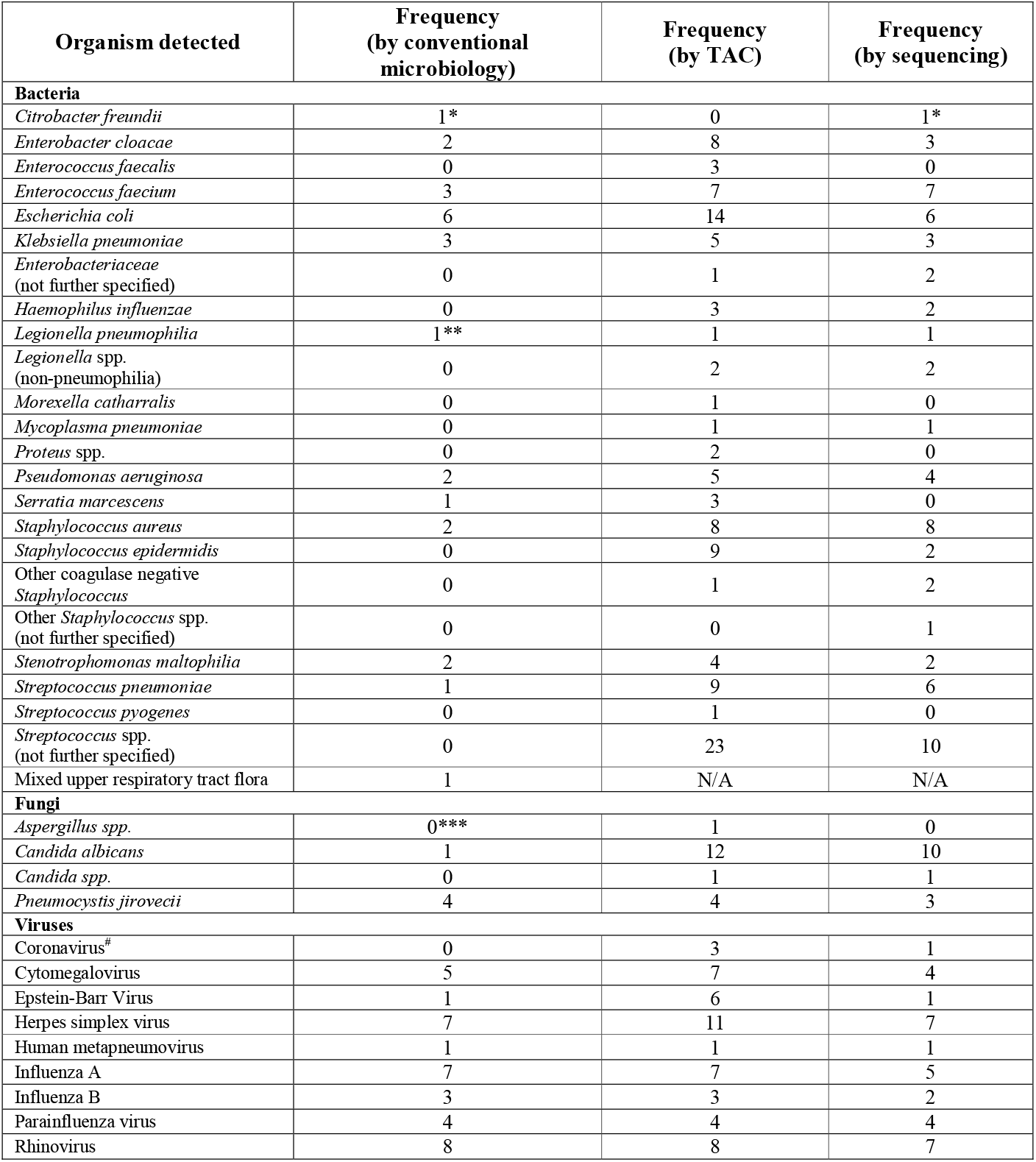
Summary of organisms detected by conventional microbiological testing (left hand column), by TAC (middle column), and by microbial sequencing (right hand column). * One hit not found in same patient; not on card. **Legionella urinary antigen test positive. *** Positive BAL galactomannan enzyme immunoassay (>0.5 units) with CT consistent with fungal pneumonia and known risk factors but fungal cultures were not positive. # refers to human coronavirade OC43, 229E and NL63, no tests were undertaken for SARS-CoV2 and final testing occurred in August 2019.

| Organism detected | Frequency<br>(by conventional<br>microbiology) | Frequency<br>(by TAC) | Frequency<br>(by sequencing) |
| --- | --- | --- | --- |
| **Bacteria** |  |  |  |
| *Citrobacter freundii* | 1 * | 0 | 1 * |
| *Enterobacter cloacae* | 2 | 8 | 3 |
| *Enterococcus faecalis* | 0 | 3 | 0 |
| *Enterococcus faecium* | 3 | 7 | 7 |
| *Escherichia coli* | 6 | 14 | 6 |
| *Klebsiella pneumoniae* | 3 | 5 | 3 |
| *Enterobacteriaceae*<br>(not further specified) | 0 | 1 | 2 |
| *Haemophilus influenzae* | 0 | 3 | 2 |
| *Legionella pneumophila* | 1 ** | 1 | 1 |
| *Legionella* spp.<br>(non-pneumophila) | 0 | 2 | 2 |
| *Moraxella catharralis* | 0 | 1 | 0 |
| *Mycoplasma pneumoniae* | 0 | 1 | 1 |
| *Proteus* spp. | 0 | 2 | 0 |
| *Pseudomonas aeruginosa* | 2 | 5 | 4 |
| *Serratia marcescens* | 1 | 3 | 0 |
| *Staphylococcus aureus* | 2 | 8 | 8 |
| *Staphylococcus epidermidis* | 0 | 9 | 2 |
| Other coagulase negative<br>*Staphylococcus* | 0 | 1 | 2 |
| Other *Staphylococcus* spp.<br>(not further specified) | 0 | 0 | 1 |
| *Stenotrophomonas maltophilia* | 2 | 4 | 2 |
| *Streptococcus pneumoniae* | 1 | 9 | 6 |
| *Streptococcus pyogenes* | 0 | 1 | 0 |
| *Streptococcus* spp.<br>(not further specified) | 0 | 23 | 10 |
| Mixed upper respiratory tract flora | 1 | N/A | N/A |
| **Fungi** |  |  |  |
| *Aspergillus* spp. | 0 *** | 1 | 0 |
| *Candida albicans* | 1 | 12 | 10 |
| *Candida* spp. | 0 | 1 | 1 |
| *Pneumocystis jirovecii* | 4 | 4 | 3 |
| **Viruses** |  |  |  |
| Coronavirus<sup>#</sup> | 0 | 3 | 1 |
| Cytomegalovirus | 5 | 7 | 4 |
| Epstein-Barr Virus | 1 | 6 | 1 |
| Herpes simplex virus | 7 | 11 | 7 |
| Human metapneumovirus | 1 | 1 | 1 |
| Influenza A | 7 | 7 | 5 |
| Influenza B | 3 | 3 | 2 |
| Parainfluenza virus | 4 | 4 | 4 |
| Rhinovirus | 8 | 8 | 7 |

### Comparison by sequencing

98 samples were available for sequencing. Metagenomic sequencing revealed 107 organisms, 100 of which were also detected by TAC (Tables 3, S3).

Concerning the 10 organisms detected by conventional microbiology or sequencing but missed by TAC, one organism, that was positive by both culture and sequencing albeit in different patients, was *Citrobacter freundii*, for which we did not have a sequence on the card. A further five pathogens were detected by sequencing (*Staphylococcus aureus, Legionella* spp., and *Staphylococcus epidermidis)* or both culture and sequencing (two *E. faecium*). Although these five were detected by TAC, they did not pass the internal quality control standards required for reporting and were considered ‘negative’ results. The remaining three organisms, two rhinovirus by conventional PCR and one *Staphylococcus* spp. by sequencing, were not detected by TAC at all.

One case of *Aspergillus fumigatus* was detected on the TAC, and although no moulds were cultured, the lavage galactomannan antigen test was highly positive (5.92 optical density index (ODI), laboratory reference range <0.5 ODI).

### Quantitation

Twenty-five organisms were grown on conventional culture at ≥10^4^ CFU/ml, the conventional cut off for quantitative culture of lavage.^20, 21^ The median cycles to threshold (Ct) for these organisms on the TAC was 27 (IQR 24-29, range 20-33). In contrast, culturable organisms detected on TAC but not on culture had a median Ct of 32 (IQR 30-34, range 22-38) (supplemental Figure S2).

### Antibiotic prescribing

Patients in the TAC and comparator cohorts had similar severity of illness, severity of respiratory failure and demographic features (Table 2). Patients managed with the TAC had a significantly different distribution of AFDs to the comparator group in the 7 days following bronchoscopy (p=0.02 by Mann-Whitney U-test), with more AFDs in the TAC cohort. This difference did not retain significance over 28 days (Supplemental Figure S5). Overall 72 (76%) of TAC patients had their antibiotics changed in the seven days following bronchoscopy, with a total of 116 changes made (Table 4). In the comparator group 50 (70%) of patients experienced a total of 65 changes. Whilst 63% of decisions in the TAC group led to de-escalation, only 37% of decisions in the comparator group were de-escalation decisions (OR 2.9 (95% CI 1.5-5.5) p=0.008 by Chi-squared). Decisions which were judged to be related to the TAC result were weighted further towards de-escalation (73% of all TAC-related changes, Table 4). 11 (30%) of escalations in the TAC group were judged to have been targeted escalations in response to TAC results. In a further six cases negative TAC results prompted investigation for alternative diagnoses.

**Table 4:** Detail of changes in antibiotic therapy in the seven days following lavage in the TAC and comparator cohorts. Changes judged to be TAC-related are shown in the left-hand sub-column for the TAC group. Several patients had more than one change in antibiotic therapy. *includes two de-escalations to prophylactic dose, ** includes two escalations from prophylactic to therapeutic dose.

| Antibiotic change | Details of change | TAC cohort |  | Comparator cohort |
| --- | --- | --- | --- | --- |
|  |  | TAC-related | TAC-unrelated |  |
| **De-escalation** | Stopping macrolides | 14 | 3 | 6 |
|  | Stopping carbapenem or anti-pseudomonal penicillin | 10 | 14 | 10 |
|  | Narrowing from carbapenem/antipseudomonal penicillin to narrower spectrum penicillin | 7 | 0 | 1 |
|  | Stopping cotrimoxazole | 6* | 1 | 0 |
|  | Stopping antivirals | 3 | 0 | 0 |
|  | Stopping aminoglycosides | 2 | 3 | 0 |
|  | Stopping other agents | 6 | 4 | 7 |
| **Escalation** | Start antivirals | 7 | 0 | 1 |
|  | Start or broaden antifungals cover | 3 | 7** | 6 |
|  | Broadened Gram negative cover (add anti-pseudomonal penicillin, aminoglycoside or carbapenem) | 3 | 16 | 28 |
|  | Add glycopeptide | 4 | 1 | 5 |
|  | Add cover for atypical organism | 1 | 1 | 1 |

## Discussion

We demonstrate that a customised molecular diagnostic, designed to meet the needs of a specific clinical setting produced accurate results in a clinically important time-frame and was associated with an increase in antibiotic-free days relative to the comparator group in the week following investigation. Diagnostic performance was similar when assessed in stored samples from multiple centres, indicating a generalisable result.

Molecular diagnostic platforms for respiratory infection syndromes have, until recently, largely focussed on viral pathogens.^16^ However, the need to optimise antimicrobial therapy whilst limiting the over-use of these drugs has led to repeated calls for bacterial-focussed diagnostics.^16,22^ TACs have been previously reported for use in pneumonia.^18,23, 24^ However, apart from our previous report^18^ that demonstrated limited clinical impact due to restricted organism coverage, none of the other reports have included ventilated patients and were restricted to retrospective analysis of stored samples. Commercial multiple-pathogen arrays that include respiratory bacteria have recently become available. However, most of reports of their use in ventilated patients remain limited to describing diagnostic performance, reporting ‘potential’ to change antimicrobial therapy rather than impact on clinical practice^3,25,26^. Concerns have been raised about the risks of over-treatment from molecular diagnostics^16,27,28^, whilst conversely promising tests with the potential to change therapy have not always proven this in clinical practice^15,18^. These commercially available assays lack the broad coverage and customisability of the TAC, with consistent concerns raised around limited organism coverage adversely impacting treatment decisions.^3,18,25,26^

Although there is now widespread acceptance of the presence of a respiratory microbiome,^29,30^ the lungs of ventilated patients present a challenge to highly sensitivemolecular diagnostics^16,^. The proximal respiratory tract of ventilated patients becomes rapidly colonised with predominantly Gram negative organisms.^31,32^ This can occur in the absence of infection, and there is a risk that highly sensitive techniques will detect colonising organisms, driving unintended increases in antimicrobial use.^16^ The use of protected lower airway specimens, with growth ≥10^4^ CFU/ml for BAL have been used to distinguish infection from colonisation.^21,33^ We adapted this approach in this study, using the quasi-quantitative Ct value provided by RT-PCR and testing protected bronchoalveolar samples. Using the comparison of the Ct values of organisms detected by culture and those detected by TAC without culture, we suggest that a Ct threshold of 32 be used to identify infecting rather than colonising organism (supplemental Figures S2, S6).

One of the problems that has beset bacterial diagnostics studies has been the absence of a ‘gold standard’ against which the candidate can be assessed,^16,22,34^ as conventional culture is imperfect. For this study we used a combination of conventional microbiology (culture and viral PCR) and metagenomic sequencing. 10 organisms identified by conventional microbiology or sequencing were not ‘detected by the TAC. Overall the TAC detected more organisms than either culture or sequencing, reflecting the higher sensitivity of qPCR. However, without a perfect validation method we cannot be certain these were not ‘false positives’ and have counted them as such for the calculation of specificity. The sequencing and culture results give clinicians considerable confidence in the results provided.

The selection of organisms targeted on the card was crucial, and informed by our previous experience where omission of key organisms significantly limited the impact of a similar card.^18^ Given the case mix of our unit, with a high proportion of immunosuppressed patients, we opted to include a number of low pathogenicity organisms, (i.e. coagulase-negative *Staphylococci(CNS), Enterococci* and *Candida albicans*), as well as Herpesviridae, which we routinely tested for. The detection of these organisms can be challenging to interpret^35^, given that many critically ill patients have a degree of immunoparesis, even if not classically immunosuppressed, ^6,36^ their significance remains uncertain. As our laboratory routinely reported these organisms on conventional microbiology the clinical team were already confronted with this issue. The inclusion of CNS also aids with the interpretation of the detection of the mecA gene, which is commonly carried by these organisms, thus helping identify MRSA. The lack of CNS on commercial cards has been noted to impair interpretation of mecA in clinical samples^25,37^. However the ready customisability of the TAC would allow units to remove such organisms, as well as add other organisms of local significance as we have done subsequently during the COVID-19 pandemic^38^.

The use of a contemporaneous comparator cohort allowed for comparisons of antibiotic prescribing within the context of the implementation of the TAC and any heightened awareness of antimicrobial stewardship it may have engendered. Despite this, the comparator cohort saw a greater proportion of escalation decisions in the week following lavage, and had fewer antibiotic-free days. The lack of difference in AFDs at day 28 is unsurprising, as suspected pneumonia is only one of multiple drivers of antibiotic use. Although the comparator and TAC groups had similar characteristics, our observational design means that we cannot be certain that unmeasured confounders did not contribute to the effects seen.

This study established a molecular diagnostic test to meet the needs of a particular intensive care unit, and implemented it in the context of a well-established antimicrobial stewardship program. Although the demonstration of similar performance on stored samples from multiple centres is reassuring, the impact on antimicrobial stewardship is likely to be context-dependent. Replication in additional settings with distinct approaches to stewardship is required before we can be certain of its external generalisability, whilst evaluation in a randomised, controlled trial would help reduce any bias that may have arisen from our observational study design. We believe this approach represents a promising new approach to the management of severe pneumonia.

## Supporting information

Supplemental methods, results, tables and figures

## Data Availability

De-identified data is available on request to the corresponding author. All samples were sequenced at the Wellcome Sanger Institute, and raw read data is available at the European Nucleotide Archive (ENA) with study accession numbers ERP111277, ERP111280, ERP112277, and ERP018622.

## Acknowledgements

The VAPrapid investigators are Prof DF McAuley, Prof TS Walsh, Dr N Anderson, Dr S Singh, Prof P Dark, Dr A Roy, Prof GD Perkins, Ms L Emerson, Prof B Blackwood, Dr SE Wright, D K Kefala, Prof CM O’Kane, Dr SV Baudouin, Dr RL Patterson, Dr A Agus, Dr J Bannard-Smith, Dr NM Robbin, Prof ID Welters, Dr C Bassford, Dr B Yates, Dr C Spencer, Dr SK Laha, Dr J Hulme, Prof S Bonner, Dr V Linnett, Dr J Sonsken, Dr Van Den Broeck, Dr G Boschman, Mr DWJ Kennan, Dr AJ Allan, Mr G Phair, Ms J Parker and Dr SA Bowett.

The authors thank the consultant intensivists of the John V Farman Intensive Care Unit, Drs P Bradley, P Featherstone, S Ford, M Georgieva, A Johnston, R Mahroof, J Martin, J Preller, K Patel, C Summers, M Trivedi, J Varley, Pharmacist L Radford, the nursing and physiotherapy teams who managed the patients, and the patients and their families who consented to the study. We also thank Torsten Seemann for access to his Kraken database.

