## Supplemental methods, results, tables and figures for "Development and implementation of a customised rapid syndromic diagnostic test for severe pneumonia"

**Inclusion and exclusion**

Patients were eligible for inclusion if they were receiving invasive mechanical ventilation, and if the treating intensive care specialist (consultant) suspected pneumonia and was planning to perform diagnostic bronchoscopy. Exclusions were lack of a proxy decision maker to provide study assent, and lack of laboratory study team availability to perform the TaqMan array card assay (the laboratory study team were routinely unavailable from Friday 5pm to Monday 8am, and also sporadically unavailable due to leave). Proxy assent (nominated or personal consultee advice) was obtained prior to study inclusion, and retrospective consent was sought if capacity was regained whilst the patient remained in hospital. Patients were identified by the treating team and included prospectively and consecutively when the laboratory study team were available.

**Development and initial validation of the card**

The local microbial ecology was reviewed using previous conventional microbiological culture data from the hospital. This was supplemented by review of the literature concerning causative organisms reported in ventilator-associated and community-acquired pneumonia and the authors’ previous experience of molecular diagnostics in pneumonia.^E1-6^ Species- or genus-specific primer/probe sequences were identified by reviewing the literature for well cited and fully validated real-time PCR assays with the presumption that where possible each organism should be covered by two sequences to minimise false positive results. In the absence of a published validated assay, one was designed in-house, normally targeting a housekeeping gene in the first instance (i.e. gyrB, rpoB, ssrA, dnaJ, recN) following the guidelines set out previously.^E7^ All assays were subjected to a comprehensive *in silico* analysis for possible cross-reactions with other high priority organisms and if necessary modified accordingly to remove any cross reaction. The species covered by the card are shown in supplemental Figure S1. The card was initially validated against our large bank of DNA extracts from a diverse range of microorganisms, known positive/negative clinical specimens, and all available EQA panels from Quality Control for Molecular Diagnostics ([www.qcmd.org](http://www.qcmd.org/)). A panel of 9 synthetic control plasmids containing all our target sequences (with 20 nucleotides each side of the primer target sites also included) were generated ([www.genscript.com](http://www.genscript.com/)) and used to quality check each batch of TAC plates and determine the limit of detection of each assay. As a demonstration of clinical utility, complete concordance was achieved against the 5 organisms from the Quality Control for Molecular Diagnostics 2018 Sepsis EQA Pilot Study (*Streptococcus pneumoniae*, *Pseudomonas aeruginosa*, *Klebsiella pneumoniae*, *Enterococcus faecalis* - in transport media and blood, and *Candida albicans* – in blood only) (Table S1).

**Bronchoscopy procedure**

Bronchoscopy for both TAC and comparator patients was conducted in accordance with the unit protocol (attached below) Where samples were taken out of hours (Monday-Thursday 5pm-8am and Sunday 8am-Monday 8am), samples were stored at 4^o^C prior to processing within 24 hours, in accordance with existing laboratory procedures. Due to lack of laboratory team availability samples were not collected between Friday 3pm and Sunday 8am.

**Nucleic acid extraction**

Nucleic acid extraction from clinical samples was undertaken using the NUCLISENS easyMAG platform (Biomerieux, Marcy L’Etoile, France), in accordance with the manufacturer’s instructions. Nucleic acids were extracted from 500 µL of input sample, with a dilution of MS2 bacteriophage added pre-extraction to act as an internal extraction and inhibition control.

**TaqMan Low-Density Array**

TaqMan Array Cards (ThermoFisher, Waltham, MA) are microfluidic cards with 8 specimen loading ports that lead to 48 inter-connected wells, each of which can be preloaded with the primers and probes necessary for independent simplex PCR reactions. Following completion of specimen loading, wells are sealed to create a closed system for each reaction to occur in parallel. Our 52-pathogen TAC used a collection of in-house primers, with more than one pan-specific or type-specific primer set included to increase overall specificity for a number of pathogens (Figure S1). The card also included primers to target the endogenous control RNase P, the internal control MS2, and the known genetic marker of *Staphylococcus aureus* virulence, Panton–Valentine leukocidin (PVL). Sequences on the card are available on request.

Cards were run on the QuantStudio 7 Flex platform (ThermoFisher), following a modified version of the method previously described.^E8^ Briefly, 50 µL of each nucleic acid extract was mixed with 50 µL of TaqMan Fast Virus 1-step mastermix (ThermoFisher) and 100 µL of RNase-free water, before 98 µL was added in 2 consecutive sample loading ports covering all 96 targets. Reverse transcriptase real-time PCR was undertaken according to the following amplification protocol: 50˚C for 5 minutes, 95˚C for 20 seconds, then 45 cycles of 95˚C for 1 second followed by 60˚C for 20 seconds. Detection of a clear exponential amplification curve with a cycle threshold (CT) value ≤38 for any single gene target was reported as a positive result for the relevant pathogen.

**Conventional microbiology for prospective study**

Samples were inoculated onto a range of solid agars and incubated in both air and 5-10% CO_2_ targeting conventional respiratory tract pathogens, *Staphylococcus aureus*, Enterobacteriales and Pseudomonads. Any organism with growth >10^4^ cfu/mL was identified to species level using matrix-assisted laser desorption isonisation time of flight (MALDI TOF) mass spectrometry (Bruker Ltd, Coventry, UK). A scanty mixed growth with no predominant organism was reported as ‘mixed respiratory tract flora’ and not characterised any further. Extended culture was performed for Legionella, Nocardia, anaerobes, fungi and Mycobacterium species. Growth at <10^3^ CFU/ml was reported as negative.

A single in-house multiplex PCR assay formed the basis of conventional testing for common respiratory viruses (adenovirus, enterovirus, human metapneumovirus, influenza A virus, influenza B virus, parainfluenza virus, rhinovirus, and respiratory syncytial virus). In-house monoplex PCR assays were used for the detection of *Pneumocystis jirovecii*. *Aspergillus* spp. were tested for by culture on Sabouraud Dextrose Agar with chloramphenicol, with or without testing for the presence of galactomannan antigen in serum (serum GM) and BAL (BAL GM) by Platelia™ *Aspergillus* enzyme immunoassay (Bio-Rad Laboratories, Hercules, CA). As well as routine culture, *Legionella pneumophila* serotype 1 was tested for by the detection of antigen in urine, using the Alere BinaxNOW™ Legionella Urinary Antigen Card (ThermoFisher) with positive tests confirmed in the national reference laboratory. Conventional laboratory methods were not routinely available to detect coronaviruses.

**Metagenomic sequencing**

Residual BAL samples (average 40 mL) from 98 out of the 100 patients were used for metagenomic sequencing. Two patients with suspected containment level 3 infections were excluded. BAL was centrifuged at 500 x g for 5 minutes to separate the host cells (pellet) from the bacterial, viral and fungal pathogens (supernatant). One mL of each sample supernatant was filtered through a 0.45 µm filter and used for viral RNA and DNA extraction using a QIAamp MinElute Virus Spin kit (Qiagen), using an on-column DNase step for viral RNA. Reverse transcription and random amplification of both viral DNA and cDNA was carried out as described previously.^E9^ The remaining supernatant was centrifuged at 3220 x g for 30 minutes and the pellet was subjected to host cell depletion using MolYsis Basic5 (Molzym, Bremen, DE) followed by bacterial/fungal DNA extraction using a QIAamp DNA Mini kit (Qiagen, Hilden, DE). Half of the DNA was submitted for HiSeq 4000 shotgun metagenomic sequencing, while the other half was used to amplify the 16S V4 region using barcoded primers,^E10,E11^ and amplicons were sequenced by Illumina MiSeq sequencing. All samples were sequenced at the Wellcome Sanger Institute, and raw read data are available at the European Nucleotide Archive (ENA) with study accession numbers ERP111277, ERP111280, ERP112277, and ERP018622. Amplicon data were analysed using Qiime2 v2019.10.0.^E12^ Single-end sequences were denoised using Deblur,^E13^ and classified using a feature classifier built from the Greengenes 13_8 99% OTUs taxonomy database. For metagenomic shotgun sequence data, human reads were first removed from the data using Bowtie2 v2.3.5.^E14^ 5’Human-depleted paired reads were then classified using Kraken2 v2.0.8.^E15^ For bacterial targets, a curated bacterial database based on the Genome Taxonomy Database^E16^ was used for classification. For viral and fungal pathogens, the standard Kraken2 viral and fungal databases were used. Qiime2 and Kraken2 tabular outputs were subsequently processed in R^E17^ to calculate the proportions of reads mapping to individual taxa for each sample.

The output for each of the sequencing approaches were compared to paired negative controls and analysed for the presence of fungal, viral and bacterial reads. Fungal and viral organisms were reported if they were the dominant species and/or the read counts were above the determined background levels. Bacterial organisms that were identified by both shotgun and 16S amplicon sequencing, or present as the dominant species in shotgun sequencing above the background levels are listed in Table S3. Organisms that had low read counts, or were only identified by 16S, but were detected by TAC were included as low confidence hits (indicated in Table S3).

Ethical and regulatory approvals

The prospective study was approved by the NHS Research Ethics Service Leeds East Research Ethics Committee (17/YH/0286) Cambridge University Hospitals NHS Foundation Trust was the sponsor. The assessment of routinely collected data from the comparator group received a consent waiver and was conducted under a protocol approved by the institutional review board (A095506). VAP-Rapid^E18^ was approved by the England and Northern Ireland (13/LO/065) and Scotland (13/SS/0074) National Research Ethics Service committees and sponsored by Newcastle upon Tyne Hospitals NHS Foundation Trust.

**Supplemental results**

**Technical validation of the TAC**

To ensure cards were spotted correctly with the primers and probes in their assigned pods, nine synthetic plasmid constructs containing all the target sequences were processed to quality-check each new batch of TAC cards in a checker-board fashion. All five microorganisms from the Quality Control for Molecular Diagnostics 2018 Sepsis EQA Pilot Study were successfully detected (Table S1).

**Prospective study TAC, microbiology and sequencing results**

Individual patient results for TAC and the corresponding results from conventional microbiology and sequencing are shown in Table S3, complementing the summary level data shown in main manuscript Table 3. 2 could not be sequenced due to presence of potential containment level 3 organisms and lack of a CL3 facility in the sequencing laboratory.

**Implementation of TaqMan array**

Our experience as researchers and clinicians leads us to suggest the following approach to using the card. Where a pathogenic organism(s) is detected at Ct value of ≤32 antimicrobials can be adjusted to target the organism(s) detected, in light of known local resistance patterns and the patient's history carriage of antimicrobial-resistant organisms. Pathogenic organisms detected at a Ct of >32 are likely to be colonisers or contaminants, although detection of respiratory viruses or atypical organisms such as *Legionella spp* or *Mycoplasma pneumoniae* at higher Ct values remain significant. The use of organism abundance to distinguish colonisation from infection is an established practice in ventilated patients^E1, E19^, with a cut-off of ≥10^4^ colony forming units (CFU) commonly used. ^E1,E19^ The detection of low pathogenicity organisms needs to be interpreted in light of the patient’s known or suspected immune status. Among the immunocompromised high levels of such organisms, especially if the sole pathogen detected, should prompt treatment. In patients in whom no relevant pathogens are detected (i.e. all organisms are at low levels, low pathogenicity organisms are detected in immunocompetent hosts, or no organisms are detected at all) consideration should be given to alternative sites of infection, alternative diagnoses and where clinical suspicion of infection is low, stopping antibiotics (Figure S6).

E11. The Earth Microbiome Project (available from www.earthmicrobiome.org accessed 25/5/20)

E12. Bolyen E, Rideout JR, Dillon MR, et al. Reproducible, interactive, scalable and extensible microbiome data science using QIIME 2. *Nat Biotechnol*. 2019;37(8):852‐857.

E13. Amir A, McDonald D, Navas-Molina JA, et al. Deblur Rapidly Resolves Single-Nucleotide Community Sequence Patterns. Gilbert JA, ed. *mSystems*. 2017;2(2):759–7.

E14. Langmead B, Salzberg SL. Fast gapped-read alignment with Bowtie 2. *Nat Methods*. 2012;9(4):357-359.

E15. Wood DE, Lu J, Ben Langmead. Improved metagenomic analysis with Kraken 2. *Genome Biol*. 2019;20(1):257.

E16. Parks DH, Chuvochina M, Waite DW, et al. A standardized bacterial taxonomy based on genome phylogeny substantially revises the tree of life. *Nature Biotechnology*. 2018;36(10):996-1004.

E17. R Core Team (2014). R: A language and environment for statistical computing. R Foundation for Statistical Computing, Vienna, Austria.(available from <http://www.R-project.org/>)

E18.	Hellyer TP, McAuley DF, Walsh TS, et al. Biomarker-guided antibiotic stewardship in suspected ventilator-associated pneumonia (VAPrapid2): a randomised controlled trial and process evaluation. *Lancet Resp Med*. December 2019:1-10.

E19.	Plachouras D, Lepape A, Suetens C. ECDC definitions and methods for the surveillance of healthcare-associated infections in intensive care units. *Intensive Care Med*. 2018;44(12):2216-2218.

**Supplemental tables**

| **Sample Content** | **Matrix*** | **TaqMan array cards results (Ct Value)** |
| --- | --- | --- |
| *Streptococcus pneumoniae* | TM | S pneumoniae #1 29.602 S pneumoniae #2 28.624  Streptococcus spp #1 29.060 Streptococcus spp #2 29.027 |
| *Pseudomonas aeruginosa* | TM | P aeruginosa #1 28.806 P aeruginosa #1 28.350 |
| *Klebsiella pneumoniae* | TM | K pneumoniae #1 25.650 K pneumoniae #2 25.106  Enterobacteriaceae 26.922 |
| *Enterococcus spp* | TM | E faecalis ddl 26.641 |
| *Streptococcus pneumoniae* | Blood | S pneumoniae #1 31.353 S pneumoniae #2 30.811  Streptococcus spp #1 31.735 Streptococcus spp #2 31.952 |
| *Candida albicans* | Blood | Candida albicans 34.313 Candida spp 33.124  Fungal 18S 30.839 |
| *Pseudomonas aeruginosa* | Blood | P aeruginosa #1 28.065 P aeruginosa #1 30.647 |
| *Klebsiella pneumoniae* | Blood | K pneumoniae #1 26.770 K pneumoniae #2 25.213  Enterobacteriaceae 26.715 |
| *Enterococcus spp* | Blood | E faecalis ddl 30.431 |
| Negative | Blood | PCR Negative |

**Table S1: Performance of TaqMan array card in the Public Health England Quality Control for Molecular Diagnostics 2018 Sepsis EQA Pilot Study** (TM-transport medium, ddl- D-alanine-D-alanine ligase)

|  | Reference standard (microbiological culture) | | total |
| --- | --- | --- | --- |
| TAC | +ve | -ve |  |
| +ve | 55 | 230 | 285 |
| -ve | 2 | 3425 | 3423 |
| total | 57 | 3655 | 3712 |

**Table S2: 2x2 table for TAC vs culture for the retrospective stored sample study.** Results presented for the 29 tests on the TAC covering culturable organisms (atypical bacteria, viruses and Pneumocystis jirovecii excluded)

| **Sample** | **TaqMan_hits** | **Ct values** | **Conventional**  **Microbiology** | **Microbial**  **Sequencing** | **Total Validated** |
| --- | --- | --- | --- | --- | --- |
| 1 | Coronavirus OC43 | 28.914/25.624 |  | ✓ | ✓ |
| 2 | Legionella pneumophila | 30.75/33.226 | ✓ | ✓ | ✓ |
| 3 | Influenza B | 35.259/25.714 | ✓ |  | ✓ |
|  | *Staphylococcus aureus (unreported)* | *36.177/34.185* |  | Staph. aureus |  |
| 4 | Influenza A | 25.969 | ✓ | ✓ | ✓ |
|  | *Influenza A H3** | *25.714* | *✓* |  | *✓* |
|  | Staphylococcus aureus | 26.829/27.302 | ✓ | ✓ | ✓ |
| 5 | Escherichia coli | 34.711/36.142 |  |  |  |
|  | Enterobacteriaceae* | 35.102 |  |  |  |
| 6 | negative | - | ✓ | ✓ | ✓ |
| 7 | Coagulase Negative Staph* | 33.536 |  | ✓ | ✓ |
|  | Staphylococcus epidermidis | 34.469 |  | ✓ | ✓ |
|  | *MecA*** | *30.995* |  |  |  |
| 8 | negative | - | ✓ | ✓ | ✓ |
| 9 | Coronavirus NL63 | 38.115 |  |  |  |
| 10 | Rhinovirus | 28.447/32.301 | ✓ | ✓ | ✓ |
|  | Influenza B | 30.274/29.312 | ✓ | ✓ | ✓ |
|  | Klebsiella pneumoniae | 28.097/28.871 | ✓ | ✓ | ✓ |
|  | Enterobacteriaceae* | 28.025 |  | ✓ | ✓ |
|  | Enterococcus faecium | 33.835/33.564 |  | (✓) | ✓ |
| 11 | Streptococcus pneumoniae | 29.48/27.47 |  | ✓ | ✓ |
| 12 | Cytomegalovirus | 29.516 | ✓ | ✓ | ✓ |
|  | Pneumocystis jirovecii | 33.628 | ✓ | ✓ | ✓ |
|  | Epstein Barr Virus | 35.57 |  |  |  |
|  | Rhinovirus | 38.933 |  |  |  |
| 13 | Candida albicans | 31.653 | ✓ | ✓ | ✓ |
|  | Candida spp* | 29.997 |  | ✓ | ✓ |
| 14 | Pneumocystis jirovecii | 28.901 | ✓ | ✓ | ✓ |
| 15 | Influenza B | 27.081/26.365 | ✓ | ✓ | ✓ |
|  | Cytomegalovirus | 35.85 |  |  |  |
|  | *Enterococcus faecium (unreported)* | *33.746/33.697* | ✓ | ✓ |  |
| 16 | Coagulase Negative Staph* | 30.462 |  | ✓ | ✓ |
|  | Staphylococcus epidermidis | 28.874 |  |  |  |
|  | *MecA*** | *30.331* |  |  |  |
|  | Streptococcus pneumoniae | 31.451 |  |  |  |
|  | Streptococcus spp* | 28.021/28.858 |  |  |  |
| 17 | Negative | - | ✓ | ✓ | ✓ |
| 18 | Influenza A | 36.073/36.538 | ✓ |  | ✓ |
|  | Influenza A H3* | 34.436 | ✓ |  | ✓ |
|  | Stenotrophomonas maltophilia | 25.179 | ✓ | ✓ | ✓ |
|  | Streptococcus pneumoniae | 35.94 |  |  |  |
| 19 | Herpes Simplex Virus | 31.549/29.524 | ✓ | ✓ | ✓ |
|  | Coagulase Negative Staph* | 31.255 |  |  |  |
|  | Staphylococcus epidermidis | 29.453 |  |  |  |
| 20 | Escherichia coli | 23.612/21.367 | ✓ | ✓ | ✓ |
|  | Enterobacteriaceae* | 22.459 |  | ✓ | ✓ |
|  | Stenotrophomonas maltophilia | 18.458 | ✓ | ✓ | ✓ |
| 21 | Streptococcus pneumoniae | 27.833/25.957 |  | ✓ | ✓ |
|  | Streptococcus spp* | 27.02/27 |  | ✓ | ✓ |
| 22 | Coagulase Negative Staph* | 31 |  | ✓ | ✓ |
|  | Staphylococcus epidermidis | 30.38 |  |  |  |
|  | *MecA*** | *25.421* |  |  |  |
|  | CMV | 34.007 | ✓ | ✓ | ✓ |
| 23 | Rhinovirus | 29.647/28.918 | ✓ | ✓ | ✓ |
| 24 | Rhinovirus | 22.857/22.678 | ✓ | ✓ | ✓ |
|  | Serratia marcescens | 30.211/32.342 | ✓ |  | ✓ |
|  | Streptococcus spp | 31.931/31.859 |  |  |  |
|  | Moraxella catarrhalis | 33.513 |  |  |  |
|  | Haemophilus influenzae | 37.097/35.339 |  |  |  |
| 25 | negative | - | ✓ | ✓ | ✓ |
| 26 | Enterobacter cloacae | 27.988/27.995 | ✓ | ✓ | ✓ |
|  | Enterobacteriaceae* | 24.71 | Citrobacter freundii | ✓ | ✓ |
|  | Serratia marcescens | 28.617 |  |  |  |
|  | Staph epidermidis | 34.237 |  |  |  |
|  | Streptococcus spp | 31.147/30.041 |  |  |  |
| 27 | Enterobacter cloacae | 27.985/29.587 | ✓ | ✓ | ✓ |
|  | Enterobacteriaceae* | 29.57 |  | ✓ | ✓ |
|  | Staphylococcus aureus | 31.504/32.666 |  | (✓) | ✓ |
|  | Klebsiella pneumoniae | 32.867/35.422 |  |  |  |
|  | Escherichia coli | 33.854/33.858 |  |  |  |
|  | Streptococcus spp | 34.628/35.956 |  |  |  |
|  | Enterococcus faecalis | 33.792 |  |  |  |
| 28 | Enterobacter cloacae | 33.815/negative |  | ✓ | ✓ |
|  | Enterobacteriaceae* | 32.809 |  | ✓ | ✓ |
|  | Escherichia coli | 35.635/33.746 |  | Citrobacter freundii |  |
|  | Enterococcus faecium | 33.823/34.716 |  | (✓) | ✓ |
| 29 | negative | - | ✓ | ✓ | ✓ |
| 30 | Herpes Simplex Virus | 24.247/21.052 | ✓ | ✓ | ✓ |
| 31 | Pneumocystis jirovecii | 34.719 | ✓ |  | ✓ |
| 32 | Herpes Simplex Virus | 27.422/26.085 | ✓ | ✓ | ✓ |
|  | Streptococcus spp | 33.859/34.864 |  |  |  |
|  | Escherichia coli | 34.092/24.28 |  |  |  |
|  | Enterobacteriaceae* | 33.079 |  |  |  |
| 33 | Enterococcus faecium | 24.697/24.763 |  | ✓ | ✓ |
|  | Escherichia coli | 25.932/24.858 | ✓ | ✓ | ✓ |
|  | Enterobacteriaceae* | 25.693 |  | ✓ | ✓ |
|  | Enterobacteriaceae proteus | 31.475 |  |  |  |
|  | Enterobacter cloacae | 32.97 |  |  |  |
|  | Candida albicans | 32.379 |  | ✓ | ✓ |
|  | Candida spp* | 28.091 |  | ✓ | ✓ |
|  | Herpes Simplex Virus | 34.709 | ✓ | ✓ | ✓ |
|  | Streptococcus sp | 34.036 |  |  |  |
|  | Coagulase Negative Staph* | 34.774 |  |  |  |
|  | Staphylococcus epidermidis | 35.19 |  |  |  |
| 34 | Candida albicans | 33.758 |  | ✓ | ✓ |
|  | Candida spp* | 30.328 |  | ✓ | ✓ |
| 35 | Legionella spp | 26 |  |  |  |
|  | Enterobacter cloacae | 32/34.407 |  |  |  |
| 36 | Enterococcus faecium | 36.332 |  |  |  |
|  | Enterococcus faecalis | 34.684 |  |  |  |
|  | Escherichia coli | 31.558/30.792 |  |  |  |
|  | Enterobacteriaceae* | 32.15 |  | (✓) | ✓ |
|  | Enterobacteriaceae proteus | 34.699 |  |  |  |
|  | Enterobacter cloacae | 31.576/36.19 |  |  |  |
|  | Candida albicans | 36.004 |  | ✓ | ✓ |
|  | Candida spp* | 32.689 |  | ✓ | ✓ |
|  | Streptococcus spp | 29.937 |  | ✓ | ✓ |
| 37 | Parainfluenza virus 3 | 35.369/41.282 | ✓ | ✓ | ✓ |
|  | Staphylococcus aureus | 30.789/30.654 |  | ✓ | ✓ |
| 38 | Herpes Simplex Virus | 31.677/29.263 | ✓ | ✓ | ✓ |
| 39 | Escherichia coli | 27.94/26.445 | ✓ | ✓ | ✓ |
|  | Enterobacteriaceae* | 23.316 |  | ✓ | ✓ |
|  | Coagulase Negative Staph | 32.182 |  |  |  |
| 40 | EBV | 24.242 | ✓ | ✓ | ✓ |
| 41 | Pseudomonas aeruginosa | 23.611/22.519 |  | ✓ | ✓ |
|  | Enterococcus faecium | 29.525/29.579 | ✓ | ✓ | ✓ |
|  | Cytomegalovirus | 35.431 |  |  |  |
|  | Aspergillus | 35.341/34.13 | raised BAL galactomannan |  |  |
| 42 | negative | - |  |  |  |
|  | *E. faecium (unreported)* | *36.189* | E. faecium | E. faecium |  |
| 43 | Herpes Simplex Virus | 33.006 |  |  |  |
| 44 | Streptococcus spp | 30.006 |  |  |  |
|  | Herpes Simplex Virus | 35.306 |  |  |  |
| 45 | negative | - | ✓ | ✓ | ✓ |
| 46 | negative | - | ✓ | ✓ | ✓ |
| 47 | Rhinovirus | 26.152/26.565 | ✓ | ✓ | ✓ |
|  | Streptococcus spp | 32.849/32.483 |  |  |  |
| 48 | Herpes Simplex Virus | 27.708/27.032 | ✓ | ✓ | ✓ |
| *49* | *Candida albicans* | *30.832* |  | *ND* |  |
|  | *Candida spp** | *27.08* |  | *ND* |  |
|  | *Staphylococcus aureus* | *34.675/34.785* |  | *ND* |  |
|  | *Epstein-Barr Virus* | *32.954* |  | *ND* |  |
|  | *Streptococcus spp* | *34.315/33.935* |  | *ND* |  |
| 50 | Klebsiella pneumoniae | 23.332/23.759 | ✓ | ✓ | ✓ |
|  | Enterobacteriaceae* | 25.06 |  | ✓ | ✓ |
|  | Candida albicans | 32.985 |  |  |  |
|  | Candida spp* | 31.006 |  |  |  |
|  | Streptococcus spp | 30.845/30.889 |  |  |  |
|  | Staphylococcus epidermidis | 35.43 |  |  |  |
|  | Stenotrophomonas maltophilia | 35.615 |  |  |  |
|  | Escherichia coli | 34.143/33 |  |  |  |
| 51 | Rhinovirus | 24/23.572 | ✓ | ✓ | ✓ |
|  | Pseudomonas aeruginosa | 27.782/27.393 |  | ✓ | ✓ |
|  | Serratia marcescens | 31/ 30.756 |  |  |  |
|  | Enterobacteriaceae | 29.107 |  |  |  |
| 52 | Streptococcus pneumoniae | 26.865/26.814 |  | ✓ | ✓ |
|  | Haemophilus influenzae | 29.252/32.253 |  | ✓ | ✓ |
|  | Streptococcus spp* | 27.818/28 |  | ✓ | ✓ |
| 53 | negative | - | ✓ |  |  |
|  | Legionella spp (unreported) | *late Ct* |  | Legionella sp |  |
| 54 | Epstein-Barr Virus | 32.612 |  |  |  |
|  | Escherichia coli | 33.168/32.146 |  |  |  |
|  | Enterobacteriaceae* | 33.3 |  | ✓ | ✓ |
|  | Streptococcus spp | 35.839 |  |  |  |
| 55 | negative | - | ✓ | ✓ | ✓ |
| 56 | negative | - | ✓ | ✓ | ✓ |
| 57 | HSV | 36.732/33.212 |  |  |  |
|  | Candida albicans | 30.441 |  | ✓ | ✓ |
|  | Candida spp* | 30.721 |  | ✓ | ✓ |
|  | Streptococcus spp | 32.424/31.792 |  | ✓ | ✓ |
| 58 | Streptococcus spp | 32.597/32.993 |  | (✓) | ✓ |
|  | Stenotrophomonas maltophilia | 34.504 |  |  |  |
|  | Cytomegalovirus | 38.142 | ✓ | ✓ | ✓ |
| 59 | negative | - | ✓ | ✓ | ✓ |
| 60 | negative | - | ✓ | ✓ | ✓ |
| 61 | Influenza A | 23.472/23.884 | ✓ | ✓ | ✓ |
|  | *Influenza A H12009** | *24.68* | *✓* |  | *✓* |
|  | Human coronavirus | 33.525 |  |  |  |
| 62 | negative | - | ✓ | ✓ | ✓ |
| 63 | Escherichia coli | 26.8/25.518 | ✓ | ✓ | ✓ |
|  | Enterobacteriaceae* | 26.842 |  | ✓ | ✓ |
|  | Streptococcus spp | 31.943/31.95 |  |  |  |
| 64 | Influenza A | 31.786/33.092 | ✓ | ✓ | ✓ |
|  | *Influenza A H12009** | *34.195* | ✓ |  | ✓ |
|  | Staphylococcus aureus | 32.593/33.529 | ✓ | ✓ | ✓ |
|  | Streptococcus spp | 30.185/31.04 |  | (✓) | ✓ |
|  | Epstein-Barr Virus | 35 |  |  |  |
| 65 | Haemophilus influenzae | 29.796/31.514 |  | ✓ | ✓ |
|  | Staphylococcus aureus | 30.433/30.241 |  | ✓ | ✓ |
|  | Streptococcus pneumoniae | 33.693/34.986 |  |  |  |
|  | Streptococcus spp* | 33.271/34.49 |  |  |  |
| 66 | Influenza A | 24.032/24.258 | ✓ | ✓ | ✓ |
|  | *Influenza A H12009** | *25.498* | *✓* |  | *✓* |
|  | Staphylococcus aureus | 31.403/31.307 |  | ✓ | ✓ |
| 67 | Influenza A | 24.553/25.573 | ✓ | ✓ | ✓ |
|  | *Influenza A H12009** | *27.372* | ✓ |  | ✓ |
| 68 | Escherichia coli | 28.879/26.58 | ✓ | ✓ | ✓ |
|  | *Enterobacteriaceae** | *27.3* |  | ✓ | ✓ |
| 69 | Pseudomonas aeruginosa | 22.442/21.451 | ✓ | ✓ | ✓ |
| 70 | negative | - | ✓ | ✓ | ✓ |
| *71* | Influenza A | 19.057/18.981 | *✓* | *ND* | ✓ |
|  | *Influenza A H12009** | *19.359* |  | *ND* |  |
|  | Staphylococcus epidermidis | 30.847 |  | *ND* |  |
|  | Coagulase Negative Staph* | 28.31 |  | *ND* |  |
|  | *Mec A*** | *30.342* |  | *ND* |  |
| 72 | Klebsiella pneumoniae | 30.116/30.92 | ✓ | ✓ | ✓ |
|  | *Enterobacteriaceae** | *31.642* |  | ✓ | ✓ |
|  | Enterococcus faecium | 29.631/34.138 |  |  |  |
|  | Streptococcus spp | 31.071/30.46 |  | ✓ | ✓ |
|  | Candida albicans | 30.627 |  | ✓ | ✓ |
|  | Candida spp* | 28.094 |  | ✓ | ✓ |
| 73 | Streptococcus pneumoniae | 22.937/20.722 | ✓ | ✓ | ✓ |
|  | Streptococcus spp* | 21.796/21.282 |  | ✓ | ✓ |
| 74 | Candida albicans | 31.319 |  | ✓ | ✓ |
|  | Candida spp* | 28.695 |  | ✓ | ✓ |
| 75 | Candida albicans | 30.957 |  | ✓ | ✓ |
|  | Candida spp* | 28.145 |  | ✓ | ✓ |
|  | Streptococcus spp | 32.386/33.246 |  | (✓) | ✓ |
| 76 | Streptococcus pneumoniae | 27.069/25.926 |  | ✓ | ✓ |
|  | Streptococcus spp* | 28.388/27.479 |  | ✓ | ✓ |
| 77 | Klebsiella pneumoniae | 30.352/29.557 |  |  |  |
|  | *Enterobacteriaceae** | *31.192* |  | ✓ | ✓ |
|  | Enterococcus faecalis | 32.972 |  |  |  |
| 78 | Enterococcus faecium | 27.461/28,78 | mixed upper respiratory tract flora | ✓ | ✓ |
|  | Candida spp | 29.432 |  | ✓ | ✓ |
|  | Staphylococcus epidermidis | 32.993 |  |  |  |
|  | *Coagulase Negative Staph** | *35.849* |  |  |  |
|  | *Mec A** | *33.906* |  |  |  |
| 79 | negative | - | ✓ | ✓ | ✓ |
| 80 | Parainfluenza virus 3 | 23.183/20.441 | ✓ | ✓ | ✓ |
|  | Streptococcus spp | 28.126/27.987 |  | ✓ | ✓ |
| 81 | Enterobacter cloacae | 33.627/33.121 |  |  |  |
|  | Enterobacteriaceae* | 32.732 |  |  |  |
| 82 | Parainfluenza virus 3 | 32.754/32.723 | ✓ | ✓ | ✓ |
|  | Streptococcus spp | 37.39 |  | ✓ | ✓ |
| 83 | Human metapneumovirus | 20.726 | ✓ | ✓ | ✓ |
|  | *Staphylococcus epidermidis (unreported)* | *36.35* |  | Staph. epidermidis |  |
| 84 | Parainfluenza virus 3 | 29.652/29.798 | ✓ | ✓ | ✓ |
| 85 | Enterobacter cloacae | 33.48/36.535 |  |  |  |
|  | *Enterobacteriaceae** | *34.897* | Rhinovirus |  |  |
|  | Streptococcus spp | 33.522 |  |  |  |
| 86 | negative | - | ✓ | ✓ | ✓ |
| 87 | Streptococcus pneumoniae | 25.414/24.813 |  | ✓ | ✓ |
|  | Streptococcus spp* | 27.097/26.671 |  | ✓ | ✓ |
| 88 | Pneumocystis jirovecii | 27.175 | ✓ | ✓ | ✓ |
| 89 | negative | - | ✓ | ✓ | ✓ |
| 90 | negative | - | ✓ | ✓ | ✓ |
| 91 | Escherichia coli | 26.282/25.058 | ✓ | ✓ | ✓ |
|  | *Enterobacteriaceae** | *25.284* |  | ✓ | ✓ |
|  | Streptococcus spp | 29.341/29.195 |  | ✓ | ✓ |
|  | Epstein-Barr Virus | 33.843 |  |  |  |
| 92 | Candida albicans | 34.63 |  | ✓ | ✓ |
|  | Candida spp* | 31.566 |  | ✓ | ✓ |
|  | Streptococcus spp | 35.922/38.979 | Rhinovirus |  |  |
| 93 | Pseudomonas aeruginosa | 27.57/25.586 | ✓ | ✓ | ✓ |
|  | Cytomegalovirus | 31.394 | ✓ | ✓ | ✓ |
|  | Staphylococcus aureus | 34.856/35.58 |  | ✓ | ✓ |
|  | Streptococcus spp* | 35.626/33.058 |  |  |  |
|  | Streptococcus pyogenes | 34.013/34.016 |  |  |  |
|  | Escherichia coli | 30/31.904 |  |  |  |
|  | *Enterobacteriaceae** | *33.452* |  |  |  |
| 94 | Mycoplasma pneumoniae | 30.119/25.063 |  | ✓ | ✓ |
| 95 | negative | - | ✓ | Staphylococcus spp |  |
| 96 | Rhinovirus | 33.484/32.082 | ✓ | ✓ | ✓ |
|  | HSV | 31.272/29.312 | ✓ | ✓ | ✓ |
| 97 | Rhinovirus | 32.087/26.5 | ✓ | ✓ | ✓ |
|  | Streptococcus spp | 32.477/35.637 |  | ✓ | ✓ |
|  | Cytomegalovirus | 39.35 | ✓ |  |  |
| 98 | Candida albicans | 28.705 |  | ✓ | ✓ |
|  | Candida spp* | 25.94 |  | ✓ | ✓ |
|  | Pseudomonas aeruginosa | 32.878/33.959 |  |  |  |
| 99 | Legionella spp | 16.766 |  | ✓ | ✓ |
|  | Herpes Simplex Virus | 37.136 |  |  |  |
| 100 | negative | - | ✓ | ✓ | ✓ |

**Table S3: TaqMan results showing all individual target hits with Ct values and whether validated by conventional microbiology and/or microbial sequencing.** *Not included in validation numbers as duplicate at sub-species or genus level detection, **MecA was not included in validation numbers. TACman hits which did not pass the internal quality control standards required for reporting are indicated by (not reported). Samples which did not undergo sequencing indicated by *ND*. (✓) indicates low confidence hits from sequencing.

|  | Reference standard (conventional culture/PCR) | | totals |
| --- | --- | --- | --- |
| TAC | +ve | -ve |  |
| +ve | 61 | 111 | 172 |
| -ve | 5 | 4030 | 4929 |
| totals | 66 | 4141 | 4207 |

**Table S4: 2x2 table for TAC vs conventional microbiology for prospective study patients.** Results presented for the 43 organisms routinely tested for in the laboratory, excluding 93 tests for patients where conventional PCR failed internal controls.

|  | Reference standard (conventional culture/PCR) | | totals |
| --- | --- | --- | --- |
| TAC | +ve | -ve |  |
| +ve | 61 | 117 | 178 |
| -ve | 5 | 5017 | 5022 |
| Totals | 66 | 5134 | 5200 |

**Table S5: 2x2 table for TAC vs conventional microbiology for prospective study patients.** Results presented for all 52 organisms tested on the card, with missing standard tests due to conventional PCR assay failure and non-testing treated as ‘negative tests’. Sensitivity and specificity were 92% (95% CI 83-98%) and 98% (95% CI 97-98%).

**Supplemental figures**

**A**

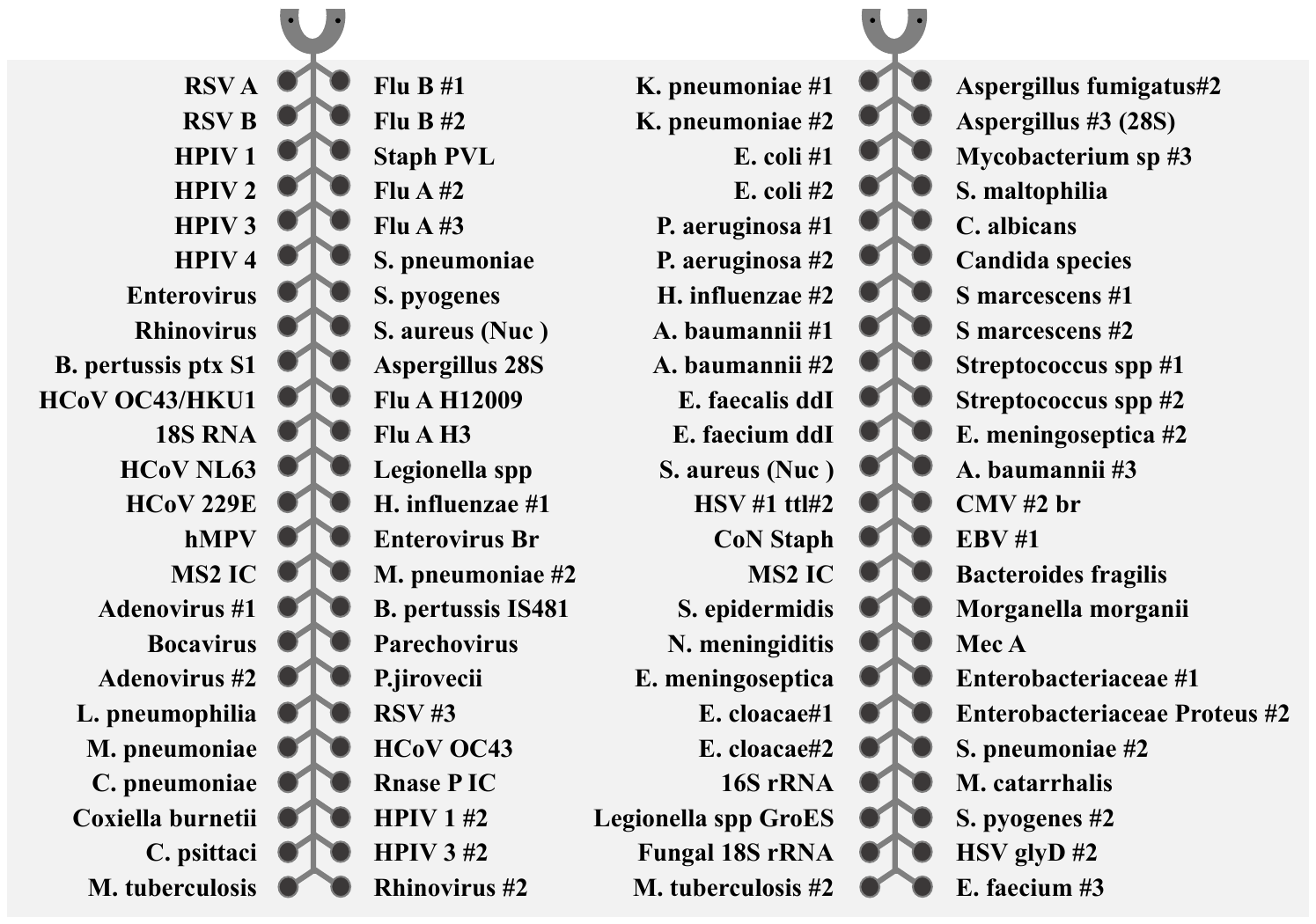

**B**

| **Bacteria** | **Mycobacteria** | **Atypical bacteria** | **Fungi** | **Viruses** |
| --- | --- | --- | --- | --- |
| A. baumannii | M. tuberculosis | C. pneumoniae* | Aspergillus fumigatus | Adenovirus |
| B. fragilis | Mycobacterium spp | C. psittaci* | Aspergillus spp | Bocavirus* |
| B. pertussis |  | C. burnetii* | Candida albicans | HCoV 229E* |
| E. cloacae |  | L. pneumophilia | Candida spp | HCoV NL63* |
| E. coli |  | Legionella spp* | Pneumocystis jirovecii | HCoV OC43* |
| E. faecium |  | M. pneumoniae* |  | Cytomegalovirus |
| E. faecalis |  |  |  | Epstein-Barr virus |
| E. meningoseptica |  |  |  | Enterovirus |
| H. influenzae |  |  |  | Herpes Simplex virus |
| K. pneumoniae |  |  |  | Influenza A |
| M. catarrhalis |  |  |  | Influenza B |
| M. morganii |  |  |  | Human Metapneumovirus |
| N. meningitidis |  |  |  | Parainfluenza |
| Proteus spp. |  |  |  | Parechovirus |
| P. aeruginosa |  |  |  | Rhinovirus |
| S. marcescens |  |  |  | Respiratory syncytial virus |
| S. aureus |  |  |  |  |
| S. epidermidis |  |  |  |  |
| Coagulase negative Staphylococci |  |  |  |  |
| S. maltophilia |  |  |  |  |
| S. pneumoniae |  |  |  |  |
| S. pyogenes |  |  |  |  |
| Streptococcus spp |  |  |  |  |

**Additional gene sequences were included for Paton Valantine Leucocidin toxin (PVL), mecA (penicillin binding protein 2a), enterobacteriaceae family and controls (MS2, 18S, RNAase P).**

**Figure S1:A) TAC layout and targets, B) summary of organisms covered.** * indicates organisms not routinely tested for by conventional microbiology in Clinical Microbiology and Public Health Laboratory, Public Health England, Cambridge. HCoV -human coronavirus.

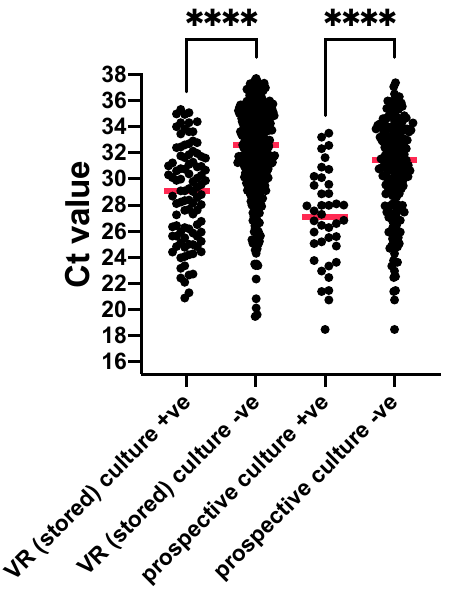

**Figure S2: Comparison of Cycles to threshold (Ct) for bacteria and fungi detected by culture and TAC (culture +ve) and those which were detected by TAC alone (culture -ve) for both the VAP-RAPID VR (stored) and prospective evaluations.** Red line indicates median value ****p<0.0001 by Mann-Whitney U test for pairwise comparisons. Organisms which are not detected by standard culture techniques were excluded.

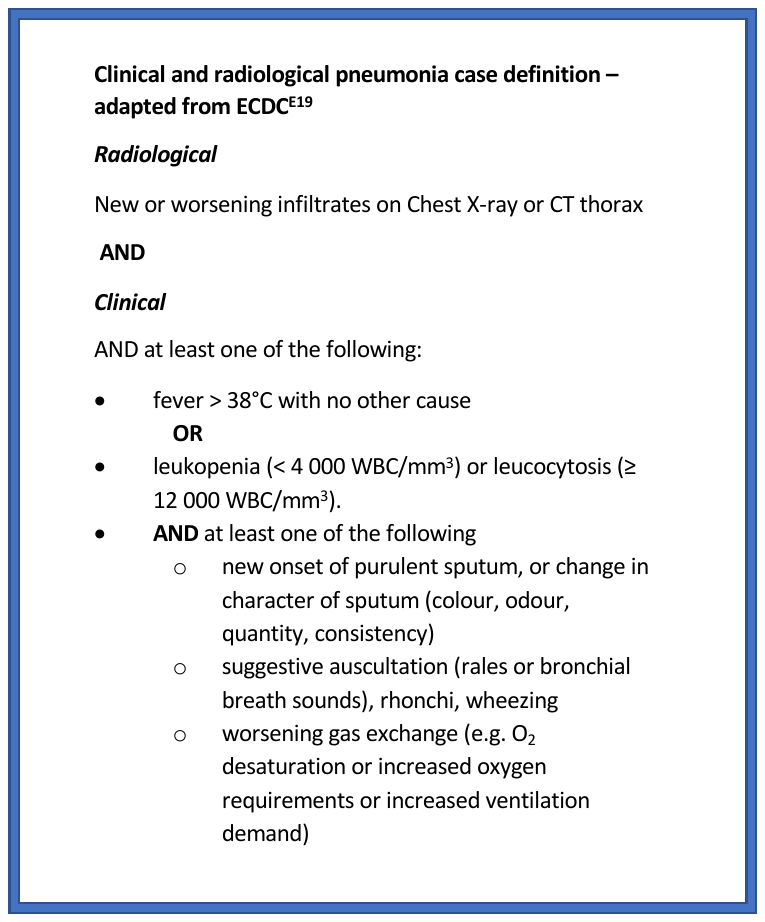

**Figure S3: clinical and radiological definition of pneumonia**

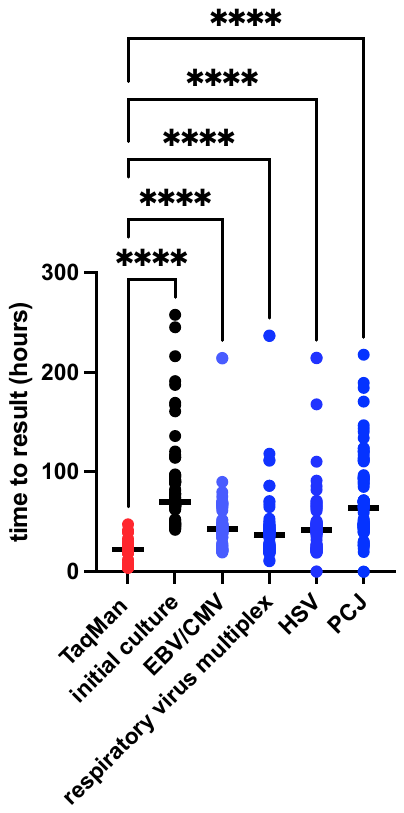

**Figure S4: Time to result for TAC, conventional culture and conventional PCR reactions.** Bold line indicates median value. Median difference in test results vs TAC was 51hrs (IQR 41-69) for culture, 21 (IQR 16-40) for Epstein-Barr virus/cytomegalovirus(EBV/CMV), 16 (IQR 5-21) for respiratory virus multiplex, 19 (IQR 15-25) for Herpes Simplex Virus (HSV) and 42 (IQR 22-70) for *Pneumocystis jirovecii* (PCJ). P<0.001 by Kruskal-Wallis, **** p<0.001 by Dunn’s post-hoc test

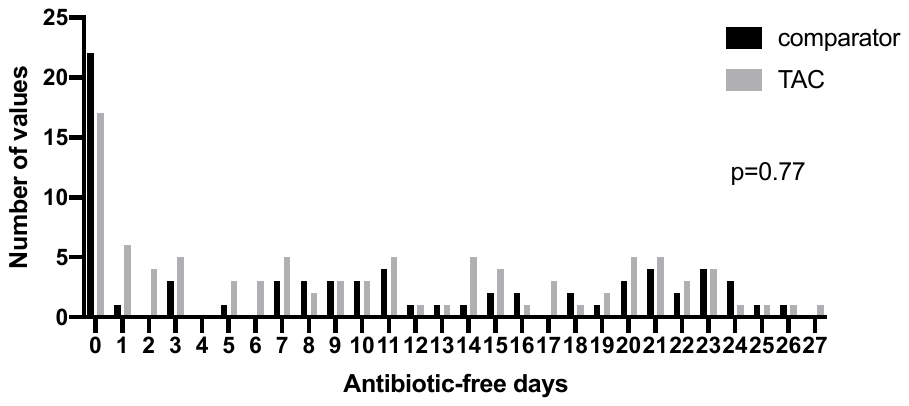

**Figure S5 Distribution of days alive and free of antibiotics in the twenty eight days following bronchoscopy and lavage in the TAC and comparator cohorts.** Following first lavage only for patients who had more than one BAL during ICU admission. P-value by Mann-Whitney U test.

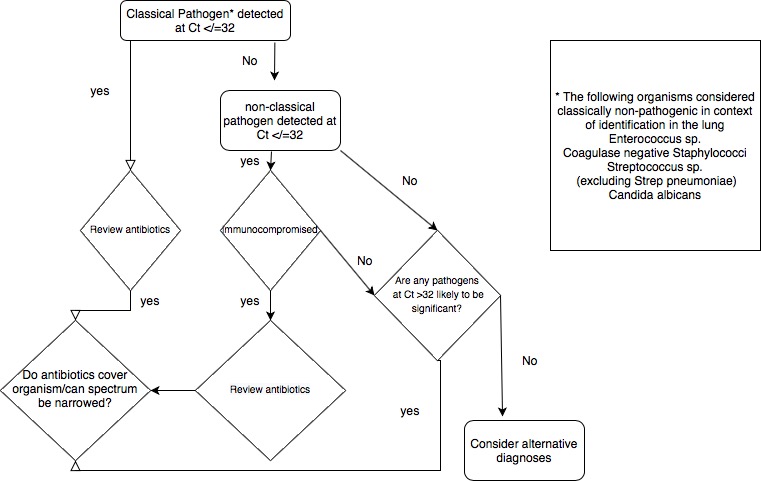

**Figure S6: Schematic representation of the proposed clinical decision tree arising from the results of the TaqMan array**.

**Rapid Pathogen Identification in Ventilated Patients with Pneumonia**

**Study protocol**

| Sponsor | Cambridge University Hospitals NHS foundation trust |
| --- | --- |
| Funder | Addenbrooke’s Charitable Trust |
| Funding Reference Number | 9366 |
| Chief Investigator | Dr Vilas Navapurkar |
| REC Number | 17/YH/0286 |
| IRAS reference | 228951 |
| Version Number and Date | Version **1.5**  **11/1/18** |

**Table of Contents**

1 Study Summary	xxxviii

1.1	Lay Summary.

1.2 Professional summary

1.3 Hypothesis

1.4 Study aims and objectives

The objectives are

1.5 Patient population

1.6 Setting

1.7 Sample size

1.8 Examination point

1.9 Assessments

2. Study Team	xli

3. Background	xliv

3.1 Background information

4. Aims and objectives	xlv

4. Study aims and objectives

The objectives are

5. Study Design	xlvi

5.4 Patients

5.4.1 Inclusion criteria

5.4.2 Exclusion criteria

5.5 Duration of study

5.6 Outcome measures

5.6.1 Primary outcome measures

5.6.2 Secondary

5.6.3 Clinical data collection

5.7 Compliance with the STARD reporting standards

6. Study procedures	xlix

6.1 Screening Procedure

6.2 Informed Consent/Assent Procedure

6.2.1	Retrospective Patient Consent

6.2.2	Withdrawal of Consent

6.4 Blood sampling

7. Study assessments	lii

7.1 Clinical Assessments

7.2 Microbiology

7.3 Additional assays

8. Collection and storage of data	liii

8.1 Recording of data

8.2 Data Management

8.3 End of Study

9. Storage of samples	liv

10. Safety	liv

11. Statistical considerations	lv

11.1 Sample Size

11.2 Data Analysis

12. Regulations, Ethics and Governance	lvi

12.1 Sponsorship

12.2 Regulatory and Ethical Approvals

12.3 Protocol Compliance

12.4 Patient Confidentiality

12.5 Good Clinical Practice (GCP)

12.6 Study Monitoring

12.7 Indemnity

12.8 Finance

12.9 Record Retention

13. Study committees	lviii

13.1 Study Management Arrangements

14. Dissemination	lviii

Abbreviations

APACHE II – Acute physiology and chronic health evaluation II (score for severity of presenting illness and allowing prediction of mortality)

CRF - Case Report Forms

CAP- community acquired pneumonia

HAP- hospital acquired pneumonia

ICU - Intensive Care Unit

ICU-AI – ICU acquired infection

IQR –interquartile range

ISF- Investigator Site File

PHE- Public Health England

PMG- project management group

PerCon-Personal consultee

ProfCon - Professional consultee

SOFA – Sequential organ failure assessment score

STARD-Standards for Reporting Diagnostic Accuracy

VAP-ventilator-acquired pneumonia

1 Study Summary

#### 1.1 Lay Summary.

Pneumonia, a serious infection of the lungs, is a common reason for Intensive Care Unit (ICU) admission. It may also develop as a significant complication of being on a mechanical ventilator. Although the clinical diagnosis is generally straight-forward, determining which organism is causing the infection (pathogen) presents a much greater challenge. Existing detection of pathogens relies on growing the organism under specific conditions in a microbiology laboratory. This process is slow, typically taking 48 to 72 hours, and is influenced by factors such as presence of antibiotics and the ease with which specific organisms can be grown. Conventional microbiology may only be positive less than 40% of cases of pneumonia ^1^, ^2^ and this means that patients are often treated with ‘best guess’ antibiotics. These antibiotics are generally broad spectrum, and risk the development of antibiotic resistance. Equally, organisms that are less commonly seen may not be covered by the initial antibiotic selection and may only be started once this organism is grown after 48 to 72 hours leading to delays in appropriate treatment. The aim of this study is to evaluate the performance of a new form of diagnostic test, using detection of pathogens by gene analysis rather than relying on growth. We believe that this approach will be more rapid and more sensitive, and therefore likely to translate into more rapid and appropriate use of antibiotics.

#### 1.2 Professional summary

Patients with severe pneumonia, both community and ventilator-acquired, present a number of challenges to clinicians. On of the most pressing issues is the rapid and accurate identification of the causative pathogen. Current practice is centered upon the use of conventional microbiological culture techniques. The requirement for microbial growth to occur means that results are frequently not available for 48 to 72 hours, and are heavily influenced by inter-current antibiotic therapy. It is not uncommon for cultures to be negative despite strong clinical evidence of pneumonia ^1^, indeed in community-acquired pneumonia this may be the case in up to 60% of cases ^1^. As are result of this, empiric, broad-spectrum antibiotic therapy is commonly employed, with the resulting risks of over-treatment, generation of antimicrobial resistance and drug-related toxicity. The aim of this study is to evaluate a new multiplex PCR test allowing for the detection of 48 relevant respiratory pathogens and resistance genes. We believe that this approach will give results within 4-6 hours, and with a much greater sensitivity, and ultimately for this to translated into more rapid, focused antibiotic treatment.

#### 1.3 Hypothesis

a) The use of a taq-man microarray plate in patients with suspected pneumonia will produce fast and accurate results compared to conventional culture.

b) The use of a taq-man microarray plate will be more sensitive for clinically significant respiratory pathogens than conventional culture.

#### 1.4 Study aims and objectives

The study aims to evaluate the diagnostic potential for a multiplex respiratory PCR based on taq-man microarray technology.

#### The objectives are

1) To develop a micro-array card for respiratory pathogens in ventilated patients ensuring broad coverage of relevant community and hospital-acquired pathogens. The card is based on existing technologies in use in the PHE Microbiology laboratory for rapid molecular diagnostics in other clinical contexts.
2) To validate the card against conventional microbiological testing using prospectively collected samples from ventilated patients with suspected pneumonia
3) To determin the speed and accuracy of the use of this card in a clinical context and the ability to return clinically useful information, relative to conventional microbiological testing.
4) To evaluate the sensitivity of the card for clinically relevant pathogens which are not detected on conventional culture.

#### 1.5 Patient population

The patient population will be ventilated patients in intensive care who are suffering clinically suspected pneumonia and who are undergoing a clinically indicated, diagnostic bronchoscopy.

#### 1.6 Setting

It is proposed to recruit patients from one general adult critical care unit within the study site. This unit all has an established research nursing teams that could be accessed for recruiting patients and collecting data. Samples will be taken to the Public Health England microbiology lab for processing and analysis.

#### 1.7 Sample size

This is an observational cohort study and we will aim to recruit 100 patients.

#### 1.8 Examination point

Samples of broncho-alveolar lavage and blood will be taken at the time of bronchoscopy for suspected pneumonia

#### 1.9 Assessments

Patients will have clinical data collected at time of enrolment, and will be followed up via their electronic health record for 7 days following sampling.

2. Study Team

| Chief Investigator (CI) | Dr Vilas Navapurkar, Consultant in Anaesthesia and Intensive Care Medicine, John V Farman Intensive Care Unit, Addenbrooke’s Hospital, Hills Road, Cambridge, CB2 0QQ |
| --- | --- |
| Principal Investigators (PIs) at Study Sites | Dr Andrew Conway Morris, Senior Research Associate and Honorary Consultant, Division of Anaesthesia, Department of Medicine, University of Cambridge, Box 93, Addenbrooke’s Hospital, Hills Road, Cambridge, CB2 0QQ  Dr Martin Curran, Consultant Clinical Scientist, Public Health England Microbiology Laboratory, Addenbrooke’s Hospital, Hills Road, Cambridge, CB2 0QQ  Dr Nick Brown, Consultant Microbiologist, Public Health England Microbiology Laboratory, Addenbrooke’s Hospital, Hills Road, Cambridge, CB2 0QQ  Dr David Enoch Consultant Microbiologist, Public Health England Microbiology Laboratory, Addenbrooke’s Hospital, Hills Road, Cambridge, CB2 0QQ  Dr Estee Torok, Honorary Consultant in Microbiology and Infectious Diseases, Department of Medicine, University of Cambridge, Addenbrooke’s Hospital, Hills Road, Cambridge, CB2 0QQ  Professor Gordon Dougan, Professor of Microbial Pathogenesis, Department of Medicine, University of Cambridge, Addenbrooke’s Hospital, Hills Road, Cambridge, CB2 0QQ  Josefin Bartholdson, Research Associate, Department of Medicine, University of Cambridge, Addenbrooke’s Hospital, Hills Road, Cambridge, CB2 0QQ  Vanessa Wong, Clinical Lecturer in Microbiology and Infectious Diseases, Department of Medicine, University of Cambridge, Addenbrooke’s Hospital, Hills Road, Cambridge, CB2 0QQ |
| Study Research Nurse | Joanne Brown |
| Study Sponsor | Cambridge University Hospitals NHS Foundation Trust |

3. Background

#### 3.1 Background information

Pneumonia is a common cause of admission to the intensive care unit, and can also develop as a secondary complication of mechanical ventilation. The diagnosis of pneumonia relies on a combination of clinical and radiographic signs, demonstrating an inflammatory infiltrate to the lung parenchyma combined with evidence of infection. The treatment is appropriate antibiotics, together with supportive care as required by the patient’s condition.

The selection of appropriate antibiotics presents a significant challenge, as between 60 and 70% of cases of pneumonia do not yield positive results on microbial cultures. This is the case in both community acquired^1^ and hospital acquired pneumonia^2^. It can take up to 72 hours for results of conventional cultures to be returned, and these two aspects mean that pneumonia is commonly treated with empiric broad spectrum antibiotics. Rapid, sensitive tests for microbes could lead to a significant reduction in antibiotic use ^1^ and use of narrower spectrum agents, which will reduce the selective pressure for anti-microbial resistant organisms.

The investigators on this study have previously shown that a multiplex polymerase chain reactions targeting respiratory pathogens can enhance the detection of such organisms in patients with community-acquired pneumonia ^1^ and immuno-compromised patients developing pneumonia ^3^. The use of a TaqMan microarray card ^3^ allows for large multiplexing of the PCR reactions, which would allow a single card to target a wide range of potential respiratory pathogens including both community-acquired and hospital-acquired organisms. It can also include additional tests, such as pan-bacterial genes (for instance 16s ^4^), allowing for the determination of bacterial load, or specific antimicrobial resistance genes ^5^.

The aim of this study is to evaluate a new TaqMan multiplex PCR array card, which targets common community and hospital-acquired respiratory pathogens. We anticipate that the results from this card will be available more rapidly than conventional culture. We also aim to evaluate the diagnostic performance of the card, compared to conventional cultures, and validate its use in the population of ventilated patients in intensive care. In addition, conventional cultures of blood have a significantly lower yield in pneumonia that respiratory samples ^6^, however as they are considerably less invasive to obtain than broncho-alveolar lavage it would be advantageous if a highly sensitive assay for bacteria could detect relevant respiratory pathogen DNA in the blood. Therefore alongside the testing of respiratory samples, we will assess the ability of the TaqMan array to detect organisms in a contemporaneously obtained blood sample.

### 4. Aims and objectives

#### 4. Study aims and objectives

The study aims to evaluate the diagnostic potential for a multiplex respiratory PCR based on taq-man microarray technology.

#### The objectives are

1) To develop a micro-array card for respiratory pathogens in ventilated patients ensuring broad coverage of relevant community and hospital-acquired pathogens. The card is based on existing technologies in use in the PHE Microbiology laboratory for rapid molecular diagnostics in other clinical contexts.
2) To validate the card against conventional microbiological testing using prospectively collected samples from ventilated patients with suspected pneumonia
3) To determin the speed and accuracy of the use of this card in a clinical context and the ability to return clinically useful information, relative to conventional microbiological testing.
4) To evaluate the sensitivity of the card for clinically relevant pathogens which are not detected on conventional culture.

5. Study Design

5.1 Study design

This is a prospective, single unit cohort observational study, with an intended recruitment of 100 patients.

5.2 Schematic recruitment diagram

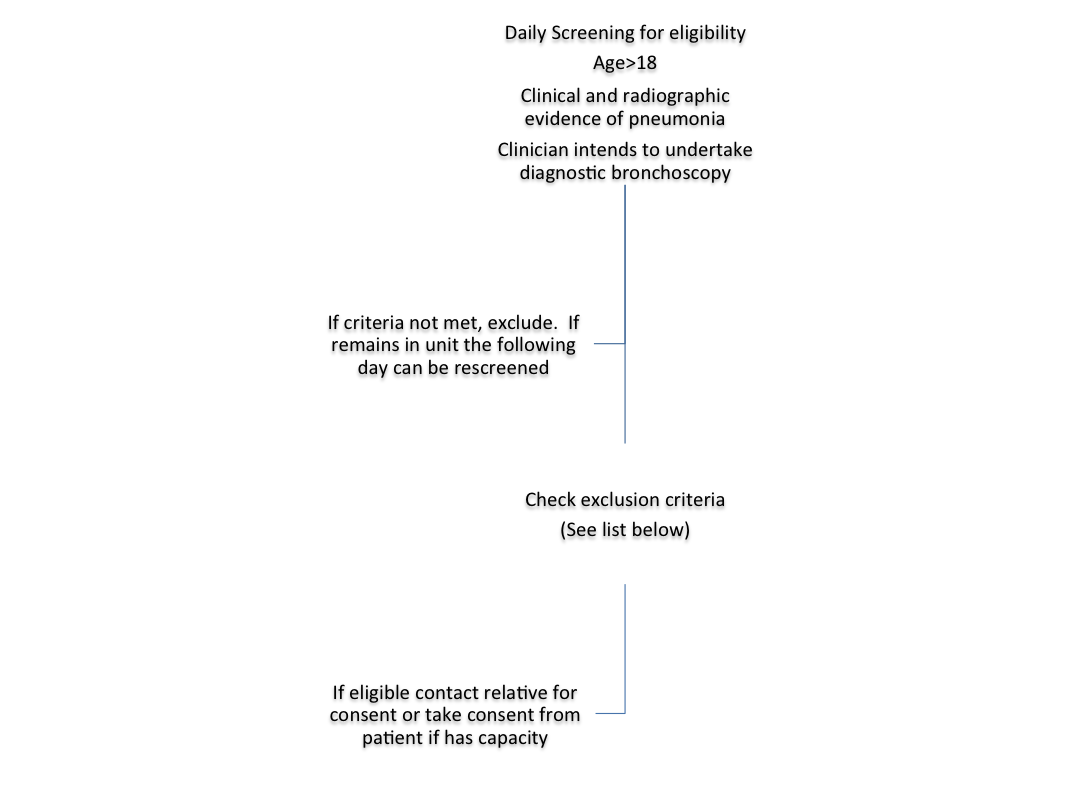
5.3 Study site

The study will recruit from a general adult critical care unit.

#### 5.4 Patients

##### 5.4.1 Inclusion criteria

- Age >18 years
- Mechanically ventilated
- Treating clinician clinically suspects pneumonia and is undertaking diagnostic bronchoscopy

##### 5.4.2 Exclusion criteria

- Lack of consent from participant
- Inability to gain advice from a personal or professional consultee

**5.4.3 Co-enrolment**

Co-enrolment will be permitted with other studies recruiting in the ICU, provided that the burden of phlebotomy is acceptable (i.e. no more than combined 50ml phlebotomy at a single time and no more than a total of 150ml across the duration of any co-enrolling studies). Co-enrolment will only be permitted with studies that have formal co-enrolment agreements with this study, and researchers must take care to not over-burden patients or proxy decision makers (Personal Consultees) with requests to take part in research studies. Co-enrolment can only occur with studies where a formal co-enrolment agreement is in place, and thus with the agreement of the chief investigators for each study.

#### 5.5 Duration of study

The study is planned to run for 3 years

#### 5.6 Outcome measures

##### 5.6.1 Primary outcome measures

The primary outcome measures are

1) Duration of time to achieve a reportable result, measured against length of time for conventional cultures to report.
2) Diagnostic performance compared to conventional diagnostic methods – with primary measures being sensitivity, specificity, positive and negative predictive value.

##### 5.6.2 Secondary

Secondary outcome measures are

1) Description of organisms detected on Taqman microarray and not detected by conventional culture.
2) Consensus expert review as to whether results would change clinical management
3) Comparison of the results of detection from broncho-alveolar lavage and blood
4) Description of clinical management following feedback of any clinically relevant results e.g. change or stop of antibiotic therapy

##### 5.6.3 Clinical data collection

Patients will be assigned a unique anonymised study code which will be used on the case record form and the study database. Routinely available clinical and laboratory data will be extracted from the electronic patient medical record (EPIC) and recorded in the case record form. This will include clinical and laboratory features of infection, routine microbiology results, antibiotic therapy, and patient outcome.

##### 5.7 Compliance with the STARD reporting standards

The analysis will be of performance of the taq-man array card compared against the current diagnostic standard of conventional culture. The reporting standards set out by Standards for Reporting of Diagnostic Accuracy (STARD) ^7^ will be adhered to (see appendix A). Data collected per the protocol will allow this to occur.

6. Study procedures

#### 6.1 Screening Procedure

Patients will be prospectively screened daily (Sunday 09.00 to Friday 14.00), on the basis of the inclusion/exclusion criteria as specified in this protocol. Screening may be performed by any qualified individual(s) designated by the PI and listed on the delegation log as having responsibility for this aspect of the study.

The site PI will be responsible for maintaining a screening log. Entries may be made by any qualified individual(s) designated by the PI. If a screened patient is not recruited the reason for not being enrolled must be recorded on the screening log.

#### 6.2 Informed Consent/Assent Procedure

Patient information sheets and informed consent forms approved by the Research Ethics Committee (REC) will be provided to the study site. The PI is responsible for ensuring that informed consent for study participation is given by each patient or a legal representative. The incapacitating nature of the condition is likely to preclude obtaining prospective informed consent from nearly all participants.

**Consent for incapacitated adults**

Advice will be sought from a personal consultee (PerCon) or nominated consultee (NomCon) should no personal consultee be available. Ordinarily the nominated consultee would be the treating consultant in Intensive Care, assuming they are not part of the study team.

Wherever possible, advice from a PerCon will be taken in person, as soon as the patient becomes eligible for inclusion. However, given the need to obtain samples in a timely fashion when infection is suspected a NomCon may provide advice if a PerCon is not available. Usual clinical practice is to start appropriate antibiotics within an hour of suspected infection, after appropriate samples have been taken.

In cases of patient incapacity, personal consultees and nominated consultees will receive a copy of the Participation Information Sheet and Consent Form. Copies will also be kept in the patient’s notes and in the Investigator Site File (ISF).

An appropriately trained doctor or nurse may take consent if delegated to do so by the local PI. If no approved form of consent is obtained a patient cannot be entered into the study.

##### 6.2.1 Retrospective Patient Consent

Patients who lack capacity at study entry will be have their capacity assessed by the clinical team. Once they regain capacity, they will be informed by the clinical team that a member of the research team would like to speak to them. A clinically qualified member of the research team (physician or research nurse) will inform the patient of their participation in the study and the circumstances of their enrolment (i.e. the advice of personal and/or nominated consultees). They will then discuss the study with the patient and he/she will be given a copy of the Patient Information Sheet to keep. The patient will be asked for consent to participate in the study. A copy of the signed Consent Form will be placed in the patient’s medical records whilst the originals will be retained by the patient and by the PI in the ISF. If the patient refuses consent, data collected about the patient will not be entered into the analysis and no further data will be collected about or from the patient and all collected and stored research samples will be destroyed. For patients who do not regain mental capacity or die by day 7, we will include them in the study based on the advice provided by the personal or nominated consultee when they were recruited to the study.

If the advice of a nominated consultee is sought, the advice of a personal consultee will be sought at the next available opportunity. If the personal consultee advises that the patient would prefer not to be involved in research, then their data and any samples obtained would be removed from the study.

##### 6.2.2 Withdrawal of Consent

Patients may withdraw or be withdrawn (by PerCon or NomCon) from the study at any time without prejudice. Data recorded up to the point of withdrawal will be included in the study analysis, unless consent to use data has also been withdrawn.

**6.3 Bronchoscopy and lavage**

Bronchoscopy and lavage will be conducted using the unit standard operating procedure (SOP) for bronchoscopy and lavage (see appendix B), and the sample processed and conveyed to the microbiology laboratory in accordance with usual unit practice. Prior to bronchoscopy, one additional step will be performed which is not routine clinical practice. The surface of the bronchoscope will be sampled with a sterile swab to allow for detection of background microbial DNA to ensure that results from the assays are not detecting environmental organisms rather than those from patients. This sample will not influence the conduct of the bronchoscopy.

Should the unit SOP for bronchoscopy be changed during the course of the study, clinicians will be expected to follow the most up to date version of the SOP and not be bound by the version that was current at the time of study initiation. Following processing for standard cultures and assays, any residual sample will be passed to the research team for the study assay.

#### 6.4 Blood sampling

Blood will be drawn in a sterile fashion, using the trust SOP for peripheral blood culture. Conventional blood cultures will be taken, in accordance with standard clinical practice for sampling in suspected pneumonia, alongside an additional research samples (**2.5ml EDTA** blood and **2.5ml P**AXgene tube).

**6.5 Clinical Management of Patients in the Study**

Clinical management of patients in this study will remain entirely at the discretion of the treating clinician. Where a clinically significant pathogen (as determined by a consultant microbiologist, independent of the study team) is detected, the results will be fed back to the responsible intensive care consultant and a decision made on treatment by the intensive care consultant in consultation with the consultant microbiologist in accordance with existing unit practice. All treatment decisions will be at the discretion of the treating intensive care consultant and enrolment in this study does not mandate or require any treatment changes.

### 7. Study assessments

#### 7.1 Clinical Assessments

Baseline data will be recorded at time of diagnostic bronchoscopy, with clinical and microbiological data recorded for the subsequent 7 days. After initial study sampling no additional sampling or clinical measurements will be mandated. Clinical data will be obtained from routinely recorded clinical information using the hospital’s electronic patient record.

##### 7.2 Microbiology

The conventional culture aliquot will be handled and reported as per the PHE-Microbiology SOPs for routine clinical samples. The same process will be undertaken for blood cultures.

From the residual BAL aliquot and EDTA blood sample, nucleic acids will be extracted and run on the Taqman microarray plate.

#### 7.3 Additional assays

Residual lavage fluid and blood and material from the pre-bronchoscopy background microbial swab will be frozen at -80°C and stored for use in additional assays, including microbial sequencing and metagenomics and patient cell gene expression studies and other relevant assays of inflammation and host response to infection.

### 8. Collection and storage of data

#### 8.1 Recording of data

All data for an individual patient will be collected by each PI or their delegated nominees and recorded in the case report form (CRF) for the study. Patient identification on the CRF will be through a unique study identifier number. A record linking the patient’s name to the unique study identifier number will be held only in a locked drawer at the study site, and is the responsibility of the PI. As such, patients cannot be identified from CRFs. Data entry will be undertaken at site by the research team. Data will be entered onto a secure study specific database which will only use the unique study number, so that patient identify will not be determinable by those without access to the linking record. Paper copies of the CRFs will remain at site and be filed in the ISF.

Data will be collected and recorded on the CRF by site research teams from the time the patient is considered for entry into the study through to the completion of outcome data. In the event that a patient is transferred to another hospital, the site research team will liaise with the receiving hospital to ensure complete data collection.

Clinical information will not be released without the written permission of the participant, except as necessary for monitoring and auditing by the Sponsor, its designee, Regulatory Authorities, or the REC. Prior written agreement from the Sponsor or its designee must be obtained for the disclosure of any said confidential information to other parties.

##### 8.2 Data Management

Data received entered into a secure database. Responsibility for maintenance of the database will rest with the chief investigator.

##### 8.3 End of Study

Recruitment will cease after recruitment of 100 patients. For regulatory and ethical reporting purposes, end of study is defined as when follow-up outcome data (7-day outcome) is collected for the last patient.

The study will stop sooner than this if mandated by the relevant REC or by the sponsor, or if funding were withdrawn.

9. Storage of samples

Samples of residual lavage fluid, nuclear material**,** serum and plasma will be labelled with the unique study identifier number and stored frozen. As described above the patient’s identity cannot be determined from the unique study identifier number alone. Samples will be retained in a secure freezer in University of Cambridge. Consent will be obtained for sample storage. Samples and data will be retained for 10 years.

### 10. Safety

Bronchoscopy is a routinely performed procedure in the intensive care unit. Although it is generally well tolerated, there are small but definite risks. These risks include transient or more prolonged impairment of oxygenation, trauma, bleeding and pneumothorax. All bronchoscopies will be undertaken for clinical indications and only residual sample remaining after conventional microbiological testing will be used in the study. The decision to undertake bronchoscopy, and assessment of the clinical risk involved, lies with the treating intensive care team and is not mandated or controlled as part of this study.

11. Statistical considerations

#### 11.1 Sample Size

We do not know what the sensitivity of the plate, in its current configuration, is and therefore a formal power calculation cannot be undertaken. This is a study to improve service development and is at the primary concept stage. The size of the study, at 100 patients, was selected to achieve a balance between cost (cards £50 each) and sufficient numbers to allow a judgment as to whether to continue to a full-scale clinical trial of the technology.

#### 11.2 Data Analysis

The primary analysis will be of the diagnostic performance of the plate compared to the existing clinical standard of quantitative culture. Using the growth of organisms at >10^4^ CFU/ml as the definition of significant growth, a well established clinical criteria^9^, the primary outcome will be the sensitivity of the diagnostic test and negative predictive value (with 95% confidence intervals) Alongside this specificity and positive predictive value will calculated (with 95% confidence intervals). The diagnostic performance of the test will be assessed by contingency table analysis, using McNemar's test to assess for the statistical significance of discordant results. Growth of organisms at >/=10^4^ colony forming units/ml of fluid on conventional will be considered the ‘current standard’ for comparison, in keeping with existing PHE laboratory practice.

Time to a reportable result will be recorded in hours, measured from the time of bronchoscopy. Difference between micro-array and conventional cultures will be assessed by median time to result for each technique, and Mann-Whitney U test of the difference to test for significance.

Secondary analysis will be of organisms that were detected by the plate but not on conventional culture, which will be handled descriptively with assessment from expert microbiologists and intensive care physicians as to the clinical relevance of the detection. Further description will be made of any organisms that were deemed to be of clinical relevance at time of detection and actions taken by the clinical team with regards to antibiotic therapy or further investigations undertaken as a result of this result. A further secondary analysis will be conducted comparing the organisms detected on broncho-alveolar lavage and from a blood sample drawn at the same time, to determin the relationship between organisms that can be detected in one or other of these body fluids.

12. Regulations, Ethics and Governance

#### 12.1 Sponsorship

The study will be sponsored by Cambridge University Hospitals NHS Foundation Trust

#### 12.2 Regulatory and Ethical Approvals

The study will be conducted in accordance with the ethical principles that have their origin in the Declaration of Helsinki. A favorable ethical opinion from a Research Ethics Committee and NHS Health Research Authority. Local R&D approval will be sought before recruitment may commence. The CI will require a written copy of local approval documentation, before sites recruit patients into the study.

#### 12.3 Protocol Compliance

The investigators will conduct the study in compliance with the protocol given favorable opinion by the REC. Changes to the protocol will require ethics committee favorable opinion prior to implementation, except when modification is needed to eliminate an immediate hazard(s) to patients. The CI in collaboration with the Sponsor will submit all protocol modifications to the REC for review in accordance with the governing regulations. Protocol compliance will be monitored by the CI who will ensure that the trial protocol is adhered to and that necessary paperwork (CRF’s, patient consent) are being completed appropriately. Any deviations from the protocol will be fully documented in source documentation and in the CRF.

#### 12.4 Patient Confidentiality

In order to maintain confidentiality, all CRFs, stored samples and study reports will identify patients by the assigned unique study identifier number only. The only link between the patient’s identity and the unique study identifier number will be held at the relevant study site, in a locked drawer).

#### 12.5 Good Clinical Practice (GCP)

The study will be carried out in accordance with the principles of the International Conference on Harmonisation Good Clinical Practice (ICH-GCP) guidelines ([www.ich.org](http://www.ich.org)). The CI, site PIs, clinical research associate and study nurse must have completed GCP training and have up to date certification before recruitment begins.

#### 12.6 Study Monitoring

12.6.1 Direct Access to Data

The agreement with the PI will include permission for study-related monitoring, audits, ethics committee review and regulatory inspections, by providing direct access to source data and study-related documentation. Consent from patients/legal representatives for direct access to data will also be obtained. Patient confidentiality will be maintained and will not be made publicly available to the extent permitted by the applicable laws and regulations.

#### 12.7 Indemnity

The participating NHS Trust has liability for clinical negligence that harms individuals toward whom they have a duty of care. Indemnity in respect of negligent harm arising from study management is provided via NHS schemes by Cambridge University Hospitals NHS Foundation Trust in its role as sponsor. Indemnity in respect of negligent harm arising from study conduct is provided by NHS schemes, via the participating NHS Trusts, covering NHS-employed staff and medical academic staff with honorary NHS contracts, who are conducting the study. Indemnity in respect of negligent harm arising from study design or protocol authorship is provided by NHS schemes, for those protocol authors whose substantive contract of employment lies with the NHS, and via University insurance schemes for protocol authors who have their substantive contract with a University. This is a non-commercial study and there are no arrangements for non-negligent compensation.

#### 12.8 Finance

This study is funded by Addenbrooke’s Charitable Trust.

#### 12.9 Record Retention

The CI and will maintain all study records according to GCP, sponsor SOP and the applicable regulatory requirements. The trial master file (TMF) will be held by the CI as per the sponsor SOP. On completion of the trial the TMF and study data will be archived by each individual site according to the applicable regulatory requirements and for up to 15 years. Following confirmation from the Sponsor, the CI will notify the PIs when they are no longer required to maintain the files. If the PI withdraws from the responsibility of keeping the study records, custody must be transferred to a person willing to accept responsibility and this must be conveyed in writing to CI.

13. Study committees

#### 13.1 Study Management Arrangements

The CI will have overall responsibility for the conduct of the study, arranging site initiation visits and providing training to site staff

- Development and distribution of the case report form and questionnaires
- Monitoring collection of data, processing data and conducting data validation.

### 14. Dissemination

The findings will be presented at national and international meetings and we aim to publish the findings in high quality peer-reviewed open access journals.

A lay person’s summary of the principal findings of the results will be sent to all patients involved in the study at their request. In addition, the most significant results will be communicated to the public through press releases.

**ICU bronchoscopy protocol**

**INTRODUCTION AND RATIONALE**

The diagnosis of a respiratory deterioration in a mechanically ventilated patient can be fraught with difficulty. There are several causes of fever and radiographic pulmonary infiltrates in patients requiring mechanical ventilation and the application of purely clinical criteria results in the over-diagnosis of ventilator associated pneumonia (VAP)^1^. Meduri *et al* confirmed the presence of lung infection in only 42% of patients with clinically suspected VAP and the frequent occurrence of non-pulmonary infections and non-infective processes ^2^.

ALI/ARDS is thought to affect 7.1% of all ICU admissions and 16.1% of all mechanically ventilated patients in European ICUs ^3^. The clinical criteria most often used for VAP diagnosis (fever, leucocytosis, purulent sputum, infiltrates on chest x-ray) are almost uniformly present in ARDS ^4^, making a clinical diagnosis of VAP extremely difficult. In addition it has been shown that, at autopsy, patients who die from ARDS are frequently found to have had a pulmonary infection that had not been recognised ^5,6^. Several trials have assessed the presence of VAP, or lower respiratory tract infection, in ARDS patients and have found that pulmonary infection affects between 34% and 70% of patients with ARDS ^7,8,9^ .

A key strategy for improving diagnostic reliability is the use of invasive sampling techniques, such as bronchoscopy. It has been shown that patients in whom bronchoscopic sampling of the lower respiratory tract is used to guide therapy have a significantly reduced mortality and less antibiotic usage than patients managed with a clinical treatment strategy ^10,11^ . Previously it has been held that blind sampling of the lower respiratory tract , such as non-bronchoscopic lavage or tracheal aspirate, is comparable to bronchoscopy, but this is questionable ^12^. While a recent study concluded that there is no difference between BAL and endotracheal aspirate the exclusion criteria of this study were such that it should not be extrapolated to the ICU patient population^13,14^.

We propose to offer a targeted bronchoscopy service to assist in the provision of high quality care to patients within the ICU who have suspected pneumonia or ALI/ARDS.

**INDICATIONS:**

All adult patients with acute respiratory failure, requiring mechanical ventilation, in association with new changes on chest radiograph, clinical suspicion of pneumonia or diagnosed ALI/ARDS.

**CONTRINDICATIONS:**

Brainstem death or likely withdrawal of treatment within 24 hours

Previous pneumonectomy

FiO_2_ requirement exceeding 0.8

Current, or recent (within 6 weeks) history of myocardial compromise (positive troponin or significant ECG changes), or a history of haemodynamically significant dysrrhythmia.

Significant coagulopathy (INR or APTTR more than 2), thrombocytopenia (platelets less than 70) or uraemia (relative contraindication).

**BRONCHOSCOPY PROTOCOL:**

1. All procedures must be undertaken in presence of a trained nurse to assist with care of the patient during the procedure ^15^.
2. A recent chest radiograph needs to be reviewed prior to the procedure.
3. Pre-oxygenation with FiO2=1.0 for at least 5 minutes.
4. The internal diameter of the endotracheal tube (ETT), through which the bronchoscope is to be inserted, must be taken into account, remembering that a standard 5.7mm bronchoscope occupies 40% of a 9mm ETT and 66% of a 7mm ETT ^15^.
5. The ventilator needs to be adjusted to a mandatory setting that will maintain minute volume and a suctionable catheter mount substituted.
6. Bronchoscopy in mechanically ventilated patients, while previously shown to be safe ^16^, should be considered a high-risk activity ^15^ and consideration given to the use of additional systemic or topical analgesia and/or neuromuscular relaxation to attempt to blunt the adverse effects of bronchoscopy on haemodynamic stability ^17^. Advice on appropriate agents, if needed, should be sought from the duty ICU Consultant prior to undertaking the procedure.
7. Avoid suction of secretions in large airways, if at all possible to prevent contamination of bronchoscope prior to collecting samples.
8. 200 ml, in aliquots, of sterile saline pre-warmed to 37°C instilled via bronchoscope, should be inserted into a sub-segmental bronchus in an area corresponding to the changes identified on chest x-ray, or in the case of bilateral diffuse shadowing in a sub-segmental division of the lingula or right middle lobe, and suctioned back into sample collection pots. The first aliquot should be discarded. Consideration should also be given to sampling from the non-affected areas.
9. The formal bronchoscopy report form (available via bluepages – “endoscopy and bronchoscopy CIS”) should be completed and a copy inserted in the patient’s medical notes. It is vital that the total volume of lavage fluid recovered is accurately documented in the report.
10. Routinely a further bronchoscopy will be undertaken 7 days after the first, or if requested by the duty ICU consultant, any other point where clinically indicated, for example, new changes identified on chest x-ray and altered clinical status.

**SAMPLE PROCESSING**

The usual recovery for a 150 ml lavage would be in the range of 40-100 ml. The sample obtained should be processed as follows:

20 ml of lavage should be sent for appropriate microbiological testing (Web OCS order code “BAL”) and any additional tests thought to be of diagnostic value in the clinical setting (immunosupression, suspected viral pneumonia, etc)
